# Neuraminidase imprinting and the age-related risk of zoonotic influenza

**DOI:** 10.1101/2025.07.03.25330844

**Authors:** Danuta M. Skowronski, Samantha E. Kaweski, Lea Separovic, Suzana Sabaiduc, Gabriel Canizares, Ayisha Khalid, Charlene Ranadheera, Nathalie Bastien, Gaston De Serres

## Abstract

Highly pathogenic avian influenza of the H5N1 subtype has shown recent unprecedented expansion in its geographic and host range, increasing the pandemic threat. The younger age of H5N1 versus H7N9 avian influenza in humans has previously been attributed to imprinted pre-immunity to hemagglutinin stalk (HA2) epitopes shared with group 1 (H1N1, H2N2) versus group 2 (H3N2) influenza A subtypes predominating in the human population before versus after 1968, respectively. Here we review the complex immuno-epidemiological interactions underpinning influenza risk assessment and extend the imprinting hypothesis to include a potential role for cross-protective neuraminidase (NA) imprinting. We compare H5N1 distributions and case fatality ratios by age and birth cohort (as proxy for HA2 and/or NA imprinting epoch) not only to H7N9 but also H5N6 and H9N2 avian influenza, representing more varied conditions of zoonotic influenza relatedness to human subtypes of the past century. We show homosubtypic NA imprinting likely further modulates the age-related risk of zoonotic H5N1 and H9N2, with implications for pandemic risk assessment and response.

---

Influenza A is a highly changeable RNA virus, error prone in its replication and with an eight-segmented genome enabling even greater diversity through reassortment of entire gene segments across viruses^1–3^. Subtypes of influenza A are designated on the basis of two surface glycoproteins encoded by gene segments 4 and 6, respectively: the hemagglutinin (HA) and neuraminidase (NA)^1^. Eighteen HA and 11 NA subtypes have been identified in nature, with all but two circulating in wild aquatic birds^2–4^. The latter comprise by far the largest natural reservoir, seeding spill-over, epi-zootics, and occasionally endemicity, among other wild and domestic animals^2–4^. The vast zoonotic pool, mutability and adaptability of influenza A viruses, and the proximity between animal hosts and human populations pose an ongoing, and recently escalating, pandemic threat.

## Pandemics and seasonal epidemics

Pandemics occur following major antigenic shift in circulating virus, notably when a novel influenza A subtype adapts to the human respiratory tract and is able to efficiently replicate and transmit from person-to-person^1,5^. Since the 20^th^ century, just three HA (H1, H2, H3) and two NA (N1, N2) subtypes of influenza A have successfully adapted to humans, causing four pandemics (1918, 1957, 1968, 2009) and one so-called pseudo-pandemic (1977)^5–9^. Each pandemic has been followed by seasonal epidemics of varying severity as descendant strains further evolved through advantageous mutation, or antigenic drift, to escape population immunity.

The 1918 H1N1 pandemic was due to an avian-origin virus that newly adapted to humans through uncertain mechanisms, with successive periods in its subsequent evolution dubbed “A swine” (“Asw”) from 1918, “A0” from 1934, and “A prime” from 1947^7,10–15^. The 1957 H2N2 pandemic was also due to an avian subtype that reassorted with the human adapted H1N1,

acquiring five of the latter’s six internal gene segments and displacing H1N1 from the human population^7,15–17^. The 1968 H3N2 pandemic was due to an avian H3 virus that reassorted with human adapted H2N2, acquiring the same five internal gene segments (originally donated by H1N1) as well the N2 surface protein^5–7,16,17^. In 1977 an H1N1 virus, phylogenetically related to the “A prime” era, re-emerged in the human population through likely unnatural means (e.g., laboratory escape), triggering a pseudo-pandemic that primarily affected H1N1-inexperienced youth <25 years^5–8,10,15^. Rather than displacing H3N2, both subtypes have co-circulated since 1977, with a human-origin H1N2 reassortment subtype also briefly but broadly circulating between 2000 and 2003^5–7,14^. During the 2009 H1N1 pandemic, a virus (called H1N1pdm09) phylogenetically related to the more distant “ASw” era re-emerged, displacing post-1977 “A prime” but not H3N2 descendant strains^5,7,9,10,13,18,19^. Older adults are widely recognized to have been relatively spared H1N1pdm09, while remaining disproportionately and severely affected by H3N2^5,7,9,10,13,18–22^.

## HA and NA interplay

While the HA is critical to viral cell entry and the NA to exit of progeny viruses, the NA also facilitates initiation of epithelial cell infection and the overall number of infected cells^2,3,23,24^. There are typically about four but up to 10 times as many HA vs. NA molecules per virion, although a more balanced 2:1 ratio has also been observed (e.g., H1N1pdm09)^2,3,25–27^.

Both proteins are targets of the antibody response but unlike HA antibodies that are virus neutralizing, NA antibodies are infection permissive while restricting replication and shedding and attenuating disease severity^28–31^. The HA and NA evolve independently to escape population immunity^6,32^; however, the more abundant and exposed HA head (HA1) is typically immunodominant and subject to greater selection pressure and evolutionary mutation than the HA stalk (HA2) or NA^2,33^.

HA subtypes are broadly categorized into two major groups based upon the phylogenetic relatedness of their more conserved HA2 segment, with greater HA2 relatedness and immunological cross-reactivity among HA subtypes belonging to the same group 1 (e.g., H1, H2, H5, H9) versus group 2 (e.g., H3, H7)^5,34–39^. Although NA subtypes are also separately categorized into group 1 (e.g., N1, N8) or group 2 (e.g., N2, N6, N9)^28^, NA antibodies are considered cross-reactive within the same (homo-subtypic) NA subtype only^40–42^. The reason some influenza A subtypes are displaced while others co-exist following pandemics likely reflects cross-reactive HA2 and/or NA effects. High pandemic attack rates induce antibodies within a large proportion of the population simultaneously; where these antibodies target shared HA2 or NA epitopes they may bind and extinguish the previously circulating subtype^43^. Such immunological interactions are more likely between H1N1 and H2N2 (HA homo-group 1) or H2N2 and H3N2 (NA homosubtypic), than between H3N2 and H1N1 (HA hetero-group and NA heterosubtypic), the latter distinction potentially explaining H3N2 and H1N1 co-circulation since 1977.

## Immunological imprinting by earliest influenza infection

Through a potent immunological phenomenon called imprinting (also known as antigenic seniority or original antigenic sin), the first influenza infection of childhood strongly influences the lifelong immune response to subsequent infecting strains^5,12,13,34,35,44–50^. Memory responses to conserved or recycled epitopes are preferentially recalled and more efficiently boosted, reinforcing swifter and more robust protection against homologous strains; conversely, de novo responses to heterologous strains may be relatively impaired^5,12,35,44,45,47^. Each new infection refocuses the memory response toward epitopes shared with the first infecting strain. Imprinting is thus a kind of immunological cohort effect induced by the earliest priming infection, but also reflects age-related accumulation of exposure and back-boosting opportunities, together recalling, reinforcing and refining pre-existing immunity (or pre-immunity) to emerging strains^5,49^.

Imprinting signatures are more readily detected within surveillance data following pandemics when exceptionally high attack rates result in widespread infection with the same or closely related virus within just a few years, particularly among highly susceptible and inter- connected children^51,52^. During the 2009 H1N1 pandemic, for example, estimated incidence approached 50% in school-aged children 5-19 years, and nearly 40% in pre-schoolers aged <5 years^20^. Despite H1N1 co-circulation since 1977, H3N2 remains the predominant human subtype overall, associated with more frequent and intense seasonal epidemics^53,54^, earlier childhood infections^35,44,55^, and on that basis most inter-pandemic imprinting and back-boosting opportunities since 1968.

## Immunological hierarchies shift in response to novel subtypes

In the context of antigenic shift, such as with novel subtype emergence or pandemics, immunological memory responses to the HA1 head become minimized, with concomitant shift in hierarchy toward more conserved HA2 and NA epitopes that can play a greater role, becoming immuno-dominant^2,5,33–35,44,49,56^. Reduced severity of the 1957 H2N2 pandemic has been attributed to such cross-protective responses to group 1 HA2 epitopes shared with earlier H1N1 imprinting^43^. The lower H1N1pdm09 risk among older adults has similarly been attributed to distant childhood imprinting to HA2 group 1 epitopes shared with post-1918 strains^10,18,19,35,57,58^.

Overall, the independent protective effects of NA pre-immunity have been rather overlooked^26,28–31,56,59–73^. On a molar basis, the NA is in fact the most immunogenic protein, evident when dissociated from the competitive HA, such as among immunologically naïve individuals or those imprinted to related NA but unrelated HA^2,25,56,64–68^. During the H2N2 period spanning 1963-1968, H2N2 vaccines induced anti-N2 responses in less than one third of recipients^26,69,74^; whereas with changed HA, H3N2 vaccines between 1968-1969 induced anti-N2 responses in at least two-thirds^69,74^. Variability in the geographic and age-related impact of the 1968 H3N2 pandemic has also been attributed to anti-N2 pre-immunity^26,28,35,43,57,60,75,76^. More recent studies reinforce the role of NA imprinting effects^28,35,70–72,77^. Pre-2009 seasonal vaccines induced anti-N1 against H1N1pdm09 that was positively correlated with older age^26,73^, likely also contributing to the attenuated H1N1pdm09 risk among older adults^20,26,78^; additionally, individuals recently infected with H1N1pdm09 raise cross-reactive anti-N1 titres against pre- 2009 seasonal strains^26,79^.

## Unprecedented H5N1 pandemic threat

Given recent unprecedented expansion in the geographic and host range of highly pathogenic avian influenza (HPAI) of the H5N1 subtype^80–83^, understanding the immuno- epidemiological intricacies of influenza has never been more important. The A/goose/Guangdong/1/1996 (Gs/GD) lineage of HPAI H5N1 was first associated with 18 human cases (6 fatal) in Hong Kong in 1997^84,85^. The Gs/GD lineage is common ancestor to all HPAI H5 viruses identified in birds or other animals since, including the nearly 1000 human cases, approximately half fatal, identified globally between 2003 and 2024^86,87^. Of note, the designation of avian influenza as high or low pathogenicity (HPAI or LPAI, respectively) indicates virulence in poultry, signalled genetically by differences in the HA cleavage site^88^, but does not predict severity in humans^89^.

In 2005, HPAI H5N1 viruses first spread inter-continentally via the flyways of migratory birds, initially evolving through diversification of the H5 gene into multiple clades and reassortment of internal gene segments with other local LPAI, notably H9N2^81,90,91^. From 2008, however, HPAI H5N1 viruses acquired an enhanced ability to reassort their NA, resulting in even greater diversification into multiple H5Nx viruses, collectively called clade 2.3.4.4^80,92^. The clade 2.3.4.4 H5N6 subtype has been endemic among birds in China and southeast Asia since 2013, with nearly 100 associated human infections between 2014 and 2024, more than half fatal^81,93^. From mid-2014, clade 2.3.4.4 viruses belonging instead to the H5N8 subtype migrated out of China, causing outbreaks in wild and domestic birds in Europe and west Asia^81^. Clade 2.3.4.4b H5N8 viruses first emerged in Egypt in 2016, seeding a further surge in outbreaks in Eurasia in 2020, including seven associated human infections, all asymptomatic, among Russian poultry workers^94–96^. Also in 2020, clade 2.3.4.4b H5N8 viruses migrated east to China where they further reassorted to acquire N6. While multiple H5N6 subclades had previously contributed, most H5N6 human cases since 2021 have been clade 2.3.4.4b^93,97^. During the 2020 European epi-zootic, H5N8 viruses also reassorted with local LPAI, resulting in clade 2.3.4.4b H5N1 viruses that spread through migratory birds to North America in late 2021^81^.

HPAI H5N1 viruses belonging to clade 2.3.4.4b have now been identified on every continent except Australia, with spillover to a remarkable range of animal hosts including wild and domestic birds, marine and terrestrial mammals, often with alarming mortality^95,98–107^.

Between March and December 2024, nearly 1000 dairy cattle herds across >15 states of the United States (US) were newly affected by clade 2.3.4.4b H5N1 viruses belonging to a unique bovine genotype (B3.13) distinct from other clade 2.3.4.4b genotypes (e.g., D1.1) affecting wild birds and poultry^105,108–110^. Sporadic human infections have followed the unprecedented global spread of clade 2.3.4.4b H5N1 viruses. Of 101 confirmed human infections due to H5N1 globally between 2021 and 2024, more than three quarters were locally acquired within Europe (n=7), North (n=68) and South America (n=2), and all with sequencing information belonged to clade 2.3.4.4b^97,111–113^. More than 90% of these cases were animal workers with only asymptomatic or mild infections, including conjunctivitis, identified through enhanced occupational awareness and/or monitoring^114,115^. Of the 67 US cases, about two-thirds (n=40) were bovine-origin.

## Imprint-based risk assessment, updated analysis inclusive of potential NA effects

While the risk of severe outcome due to influenza and other non-influenza respiratory viruses typically increases with age^116–120^, human zoonotic cases of H5N1 show an unusually youthful profile, relatively sparing older adults as highlighted earlier for H1N1pdm09^84,121,122^. This pattern of age-related risk for zoonotic H5N1 sharply contrasts with human zoonotic cases of H7N9 avian influenza which more typically and severely affect older adults^44,121,122^. Citing group-specific relatedness in the HA2 stalk relative to human influenza subtypes, and analyzing human zoonotic case and death reports through 2015, Gostic et al^44^ attributed the relative paucity of H5N1 among pre-1968 birth cohorts to cross-protective imprinting induced to HA2 group 1 epitopes shared with H1 and H2 subtypes circulating within the human population pre-1957 and 1957-67, respectively, and conversely the paucity of H7N9 among post-1968 cohorts to imprinted group 2 (H3) pre-immunity. Their hypotheses, however, focused on the HA2, disregarding the potential cross-protective effects of imprinted NA pre-immunity.

Here we expand upon earlier imprinting hypotheses to include a role not only for group-specific HA2 but also homosubtypic NA pre-immunity in modulating the age-related risk of zoonotic influenza. We quantify and compare both HA and NA relatedness among representative avian, human zoonotic, and human adapted subtypes, the latter including all publicly available sequences since 1918. Having assembled updated datasets of human zoonotic cases spanning 2003 to 2024 (**Fig. 1**), we compare H5N1 distributions and case fatality ratios (CFRs) by age and birth cohort not only to H7N9 but also H5N6 and H9N2, representing a kind of natural experiment inclusive of more varied conditions of HA2 and/or NA relatedness to human adapted subtypes of the past century. Underpinning all analyses is the assumption that, absent differential pre-immunity, age-related risks should be similar between zoonotic subtypes, with severity increasing with age.

**Fig. 1.**
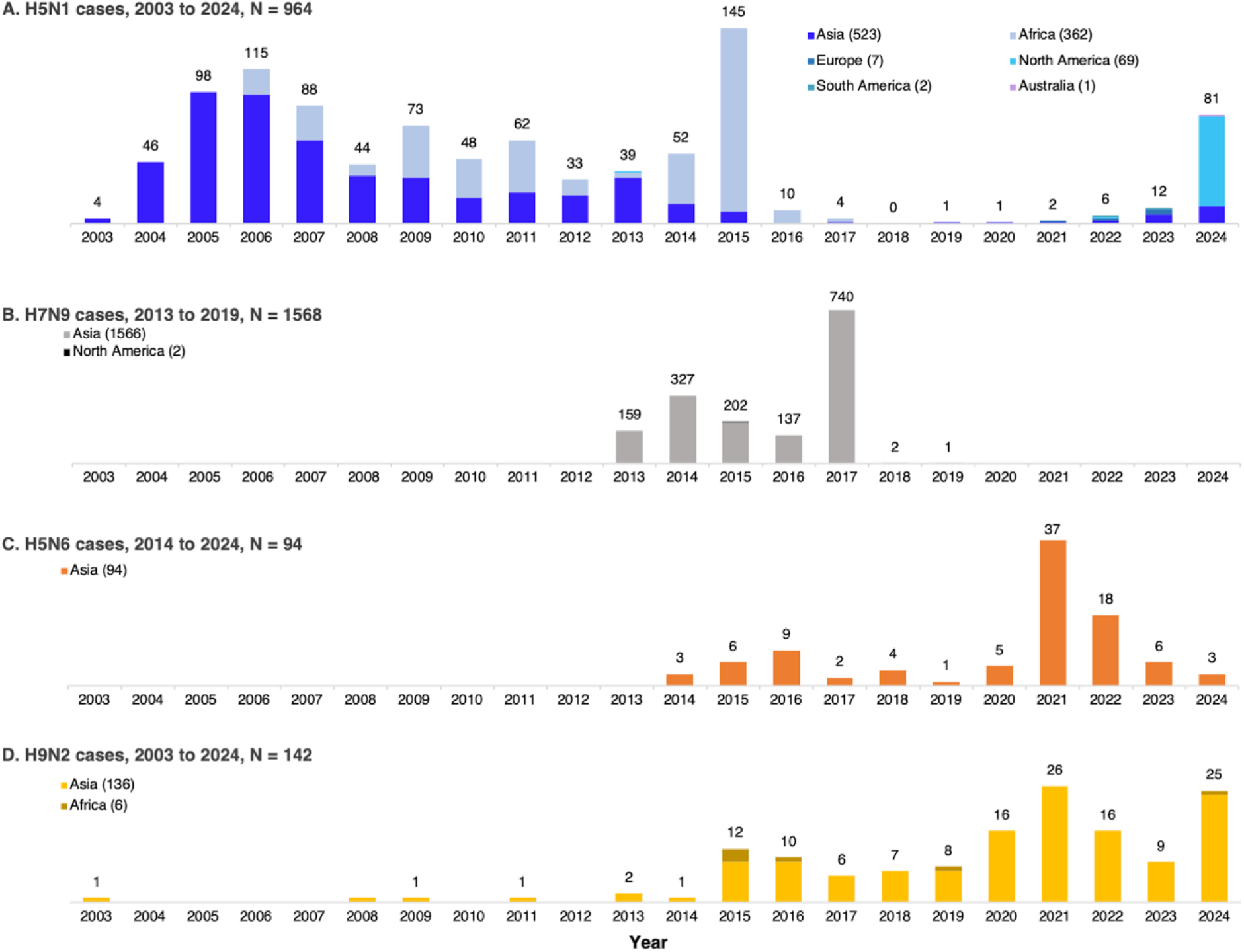
Annual human zoonotic H5N1, H7N9, H5N6 and H92 case reports, 2003-2024. Displayed are the tallies of laboratory-confirmed human zoonotic cases reported globally between 2003 and 2024 and captured within our assembled datasets regardless of completeness for age or outcome, shown by year of occurrence for (**A**) H5N1, (**B**) H7N9, (**C**) H5N6, and (**D**) H9N2. Overall tally by region is provided in parentheses within the legend for each subtype.

## HA2 and NA relatedness between zoonotic and human subtypes, 1918-2024

Broadly summarized, HPAI H5N1 belongs to HA group 1 (with H1 and H2) and is NA homosubtypic to H1N1; H7N9 (both HPAI and LPAI) instead belongs to HA group 2 (with H3) but also differs from H5N1 in being NA heterosubtypic to all human subtypes of the past century; HPAI H5N6 shares H5 with H5N1 but is, like H7N9, NA heterosubtypic to all human subtypes; and finally, LPAI H9N2 shares HA group 1 with H5, H1 and H2 while additionally being NA homosubtypic to H2N2 and H3N2. We further quantify and compare genetic relatedness of these zoonotic subtypes based upon the percentage of amino acid residues that are identical (percent identities) within matched HA1, HA2 and NA head domains (see **Methods**) (**Table 1**).

**Table 1.** Percent amino acid pairwise identities in HA1, HA2 and NA head domain of select human zoonotic and human adapted influenza A viruses. Displayed are percent amino acid pairwise identities between aligned hemagglutinin (HA) head (HA1), HA stalk (HA2), and neuraminidase (NA) head domains of human zoonotic and human adapted influenza A subtypes. Representative avian, human zoonotic and human adapted viruses included in pairwise comparisons and their precise identity values are shown in **Supplementary Tables 1 (HA1), 2 (HA2) and 3 (NA)**. **Supplementary** Figure 1 shows domain-specific relatedness observations are consistent across human adapted strains since 1918.

| Human adapted influenza A subtypes, circulation periods (HA, NA group) |  | Human zoonotic influenza A virus subtypes<br>Viral domain (group), percent amino acid pairwise identities (%) |  |  |  |  |  |  |  |  |  |  |  |
| --- | --- | --- | --- | --- | --- | --- | --- | --- | --- | --- | --- | --- | --- |
|  |  | H5N1 |  |  | H5N6 |  |  | H9N2 |  |  | H7N9 |  |  |
|  |  | HA1<br>(1) | HA2<br>(1) | NA<br>(1) | HA1<br>(1) | HA2<br>(1) | NA<br>(2) | HA1<br>(1) | HA2<br>(1) | NA<br>(2) | HA1<br>(2) | HA2<br>(2) | NA<br>(2) |
| H1N1<br>(1,1) | 1918 pandemic | 56-58 | 82-84 | 91-94 | 57 | 83 | 47 | 46 | 64 | 45 | 38 | 58-59 | 47 |
|  | 1934 – 1957 | 53-58 | 80-83 | 86-89 | 54-58 | 80-82 | 45-46 | 46-47 | 62-63 | 45-46 | 37-38 | 56-57 | 46-48 |
|  | 1977 – 2008 | 53-55 | 79-83 | 85-89 | 53-55 | 80-82 | 44-46 | 46-47 | 61-63 | 45-47 | 36-37 | 54-56 | 45-47 |
| H1N1pdm09<br>(1,1) | 2009 pandemic | 53-56 | 82-83 | 90-92 | 54 | 82 | 47 | 44 | 62 | 46 | 36 | 56-57 | 47-48 |
|  | 2015 – 2024 | 53-56 | 81-83 | 87-91 | 54-55 | 81-82 | 46 | 44-47 | 62-63 | 47 | 35-36 | 56-58 | 46-48 |
| H2N2<br>(1,2) | 1957 pandemic | 67-70 | 87-89 | 45-46 | 68 | 87 | 51 | 47 | 62 | 86 | 36-38 | 54-55 | 50 |
|  | End H2N2 epoch | 66-68 | 86-88 | 45-46 | 66 | 87 | 50 | 46 | 62 | 86 | 35-37 | 54-55 | 49 |
| H3N2<br>(2,2) | 1968 pandemic | 35-37 | 52-54 | 45-46 | 35 | 53 | 50 | 33 | 50 | 86 | 34-36 | 65-66 | 49 |
|  | 2022 | 34-36 | 50-52 | 45-46 | 34 | 51 | 49 | 33 | 49 | 81 | 34-35 | 65 | 47 |

We begin by noting that since emergence and despite evolution, HPAI H5N1 viruses are highly conserved in their HA2 and NA domains. In fact, across representative HPAI H5N1 and H5N6 avian and human zoonotic case viruses, spanning original Gs/GD through clade 2.3.4.4b, all H5 identities exceed 95% in the HA2 (versus 85% in the HA1) (**Supplementary Tables 1and 2**). Homosubtypic N1 identities among H5N1 viruses also exceed 90%, whereas heterosubtypic NA identities between H5N1 and H5N6 are <50% (**Supplementary Table 3**).

In comparing human zoonotic relative to human adapted subtypes (**Table 1**), the greatest HA2 identities are between H5 and H2 (86-89%) followed by H5 and H1 (79-83%). Of note, H5 and H2 share HA1 head identity (66-70%) comparable to the HA2 identity between H7 and H3 (65-66%), similar also to the HA2 identity between H9 and H1 or H2 (61-64%). All other HA1 and HA2 identities are <60%.

Homosubtypic NA generally exceed homo-group HA2 identities (**Table 1**). For H5N1, identity is highest relative to the N1 of 1918 (91-94%) and 2009 pandemic (90-92%) or H1N1pdm09 seasonal (87-91%) strains, less so in relation to the 1977 pseudo-pandemic virus (85-86%). For H9N2, homosubtypic N2 identities substantially exceed HA2 identities relative to pandemic or more proximal H2N2 (86%) or H3N2 (81-86%) strains. Heterosubtypic N9 and N6 vs N1 or N2 identities are again ≤50%. NA head also exceed HA1 head identities, with few exceptions similarly involving heterosubtypic NA. These domain-specific patterns summarized in relation to representative human strains in **Table 1** and **Supplementary Tables 1-3**, are shown in **Supplementary Fig. 1** to be consistent in relation to all human adapted strains with available sequences since 1918.

## Human zoonotic influenza datasets, 2003 to 2024

Absent officially curated, complete and consolidated databases with necessary case details, we constructed individual-level line-lists of virologically confirmed human zoonotic H5N1, H7N9, H5N6 and H9N2 cases as outlined in **Supplementary Notes 1-4** where we also provide completeness checks and references. As per WHO official tallies we include human zoonotic cases from 2003, and our datasets span cases occurring through December 31, 2024 (**Fig. 1**).

Our H5N1 line-list is complete for exact age in years for 872/964 (91%) cases reported from 24 countries between 2003-2024 (**Supplementary Table 4**), including 458/465 (98%) known fatalities. Of 92 (10%) cases without exact age in years, most (67; 73%) were reported from the US (one in 2022, 66 in 2024), as shown by state and avian or bovine source in **Supplementary Fig. 2**. Half the cases with known sex are female (470/876; 54%). We include 60% more H5N1 cases (872/545) and 67% more deaths (458/274) with known age than the earlier Gostic et al^44^ analysis spanning 2003-2015.

Our H7N9 line-list is complete for exact age in years for 1549/1568 (99%) cases reported from six countries (1537; 98% China) between 2013-2019, including 299/616 (49%) known fatalities. Greater missingness for the latter is due to retrospective aggregate reporting that could not be individually reconciled. About one-third with known sex are female (465/1547; 30%).

Abrupt end to the H7N9 epi-zootic in China followed poultry vaccination that began September 2017, with just 1-2 human cases thereafter in 2018 and 2019 (**Fig. 1**)^123^. Of human zoonotic H7N9 virus sequences including the HA cleavage site within GISAID (1139/1568; 73%)^124^, 96% were LPAI with just 49 (4%) HPAI, all after 2016. We include four times as many H7N9 cases (1549/355) and deaths (299/70) than the earlier Gostic et al^44^ analysis spanning 2013-2015.

Our H5N6 line-list is complete for exact age in years for all 94 cases reported by two countries (one Lao PDR, the rest China) between 2014-2024, including 47/57 (82%) known fatalities, the latter also incomplete due to aggregate reporting. About half with known sex are female (44/94; 47%). Finally, our H9N2 line-list is complete for age for 141/142 (99%) H9N2 cases reported by 9 countries (126; 89% from China) between 2003-2024, including the three known fatalities. About half with known sex are female (78/141; 55%).

## Case and fatality distributions by age and birth cohort

We examined case and fatality distributions, including CFRs, by age and birth cohort excluding those missing age in years. To address systematic differences in severe outcome ascertainment based on enhanced occupational vs. routine severe outcome surveillance we also excluded cases known to be asymptomatic and/or commercial animal workers. See **Methods** and **Supplementary Notes 1-4** for further details. Unless otherwise specified, all comparisons quantified below are statistically significant at p<0.001.

## H7N9 cases and deaths older than H5N1

Within our dataset and as in previous analyses^44,121,122^, human zoonotic H5N1 cases are substantially younger than H7N9, with median (interquartile range, IQR) of age of 18 (5-31) and 57 (45-67) years and birth year of 1990 (1978-2004) and 1959 (1948-71), respectively (**Fig. 2**), similar with consistent restriction to cases occurring since 2014 (**Fig. 3**). Individuals <18 years comprise more cases and deaths due to H5N1 (49% and 41%) than H7N9 (4% and <1%) (**Fig. 4**, **Supplementary Fig. 3**). Conversely, adults ≥50 years comprise more cases and deaths due to H7N9 (68% and 76%) than H5N1 (5% and 3%), with ≥65-year-olds contributing substantially to H7N9 (31% and 40%) but negligibly to H5N1 (1% and 1%). Age-related patterns are also similar with restriction to cases between 2013-17 (span of 99% of H7N9 cases) (**Supplementary Fig. 4**) and among fatal cases (**Fig. 4, Supplementary Fig. 3, Supplementary Fig. 5**).

**Fig. 2.**
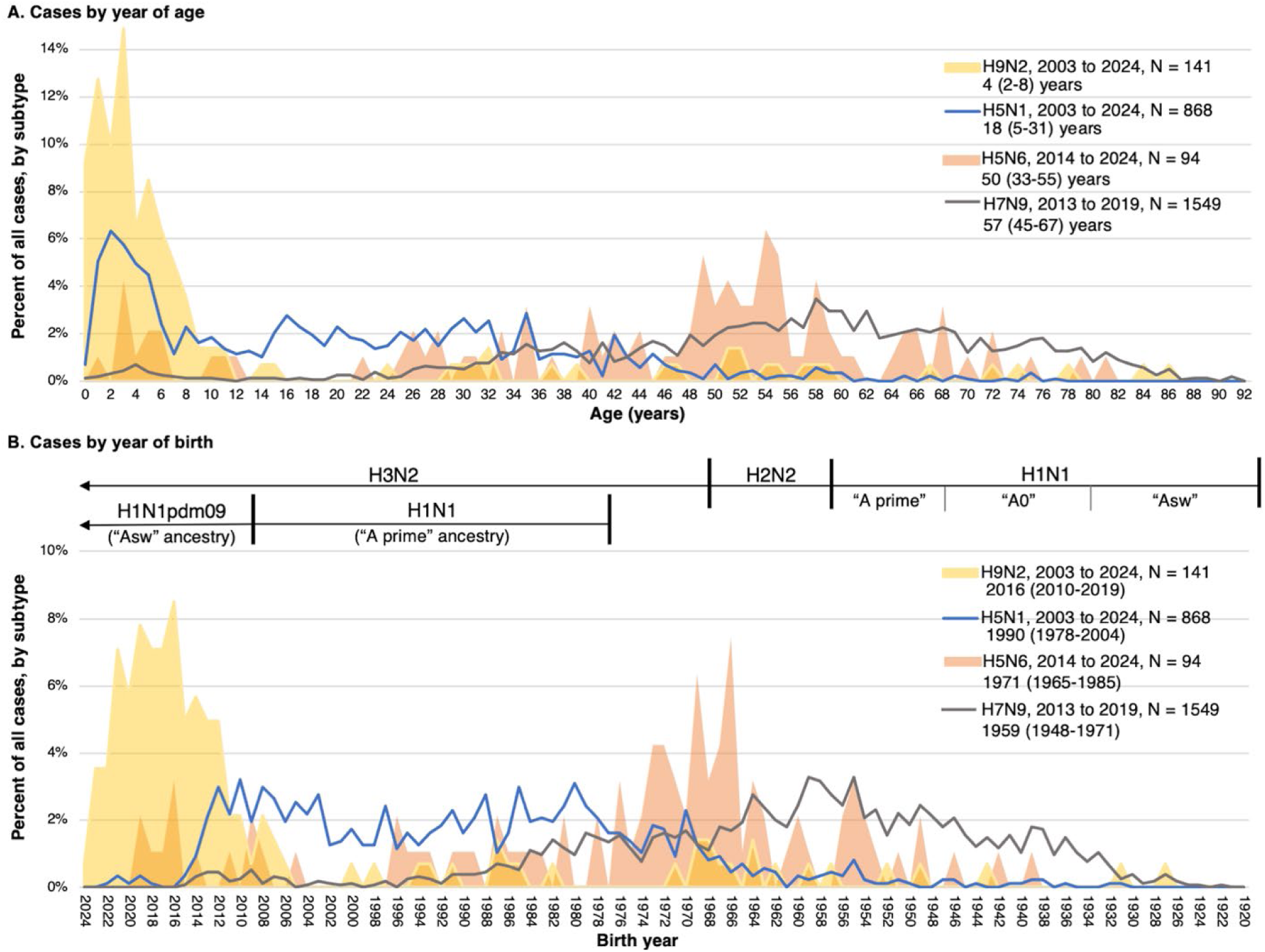
Percentage distribution of human zoonotic influenza A cases overall, by year of age and birth year. Displayed are the percentage distributions of laboratory-confirmed human zoonotic influenza A H9N2, H5N1, H5N6 and H7N9 cases with known age *overall* shown by (**A**) year of age and (**B**) birth year. Panel legends include respective span of case occurrence years, case tallies and median (interquartile range) of age in years (A) or birth years (B) for each zoonotic subtype. Displayed above panel B are the human adapted influenza A subtype(s) circulating during the corresponding birth year; the post-1918 H1N1 pandemic period is sub-divided to reflect descendant strains broadly categorized as “A swine” (“Asw”), “A0” and “A prime”, with “A prime” era virus re-emerging during the 1977 H1N1 pseudo-pandemic and “Asw” era virus re- emerging during the 2009 H1N1pdm09 pandemic.

**Fig. 3.**
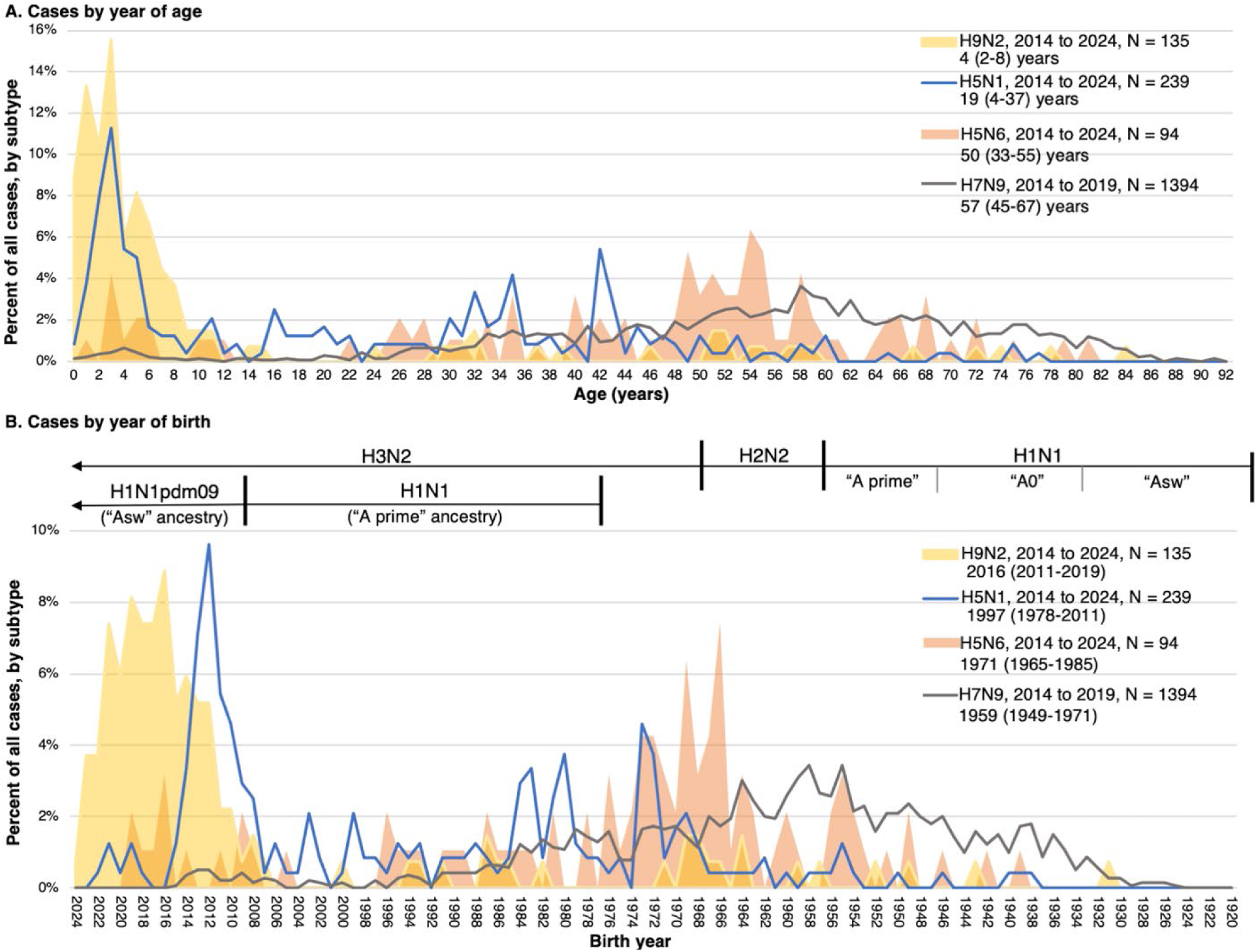
Percentage distribution of human zoonotic influenza A cases since 2014, by single year of age and birth year. Displayed are the percentage distributions of laboratory-confirmed human zoonotic influenza A H9N2, H5N1, H5N6 and H7N9 cases with known age *restricted to cases since 2014* shown by (**A**) year of age and (**B**) birth year. Panel legends include respective span of case occurrence years, case tallies and median (interquartile range) of age in years (A) or birth years (B) for each zoonotic subtype. Displayed above panel B is the human adapted influenza A subtype circulating during the corresponding birth year; the post-1918 H1N1 pandemic period is sub-divided to reflect descendant strains broadly categorized as “A swine” (“Asw”), “A0” and “A prime”, with “A prime” era virus re-emerging during the 1977 H1N1 pseudo-pandemic and “Asw” era virus re-emerging during the 2009 H1N1pdm09 pandemic.

**Fig. 4.**
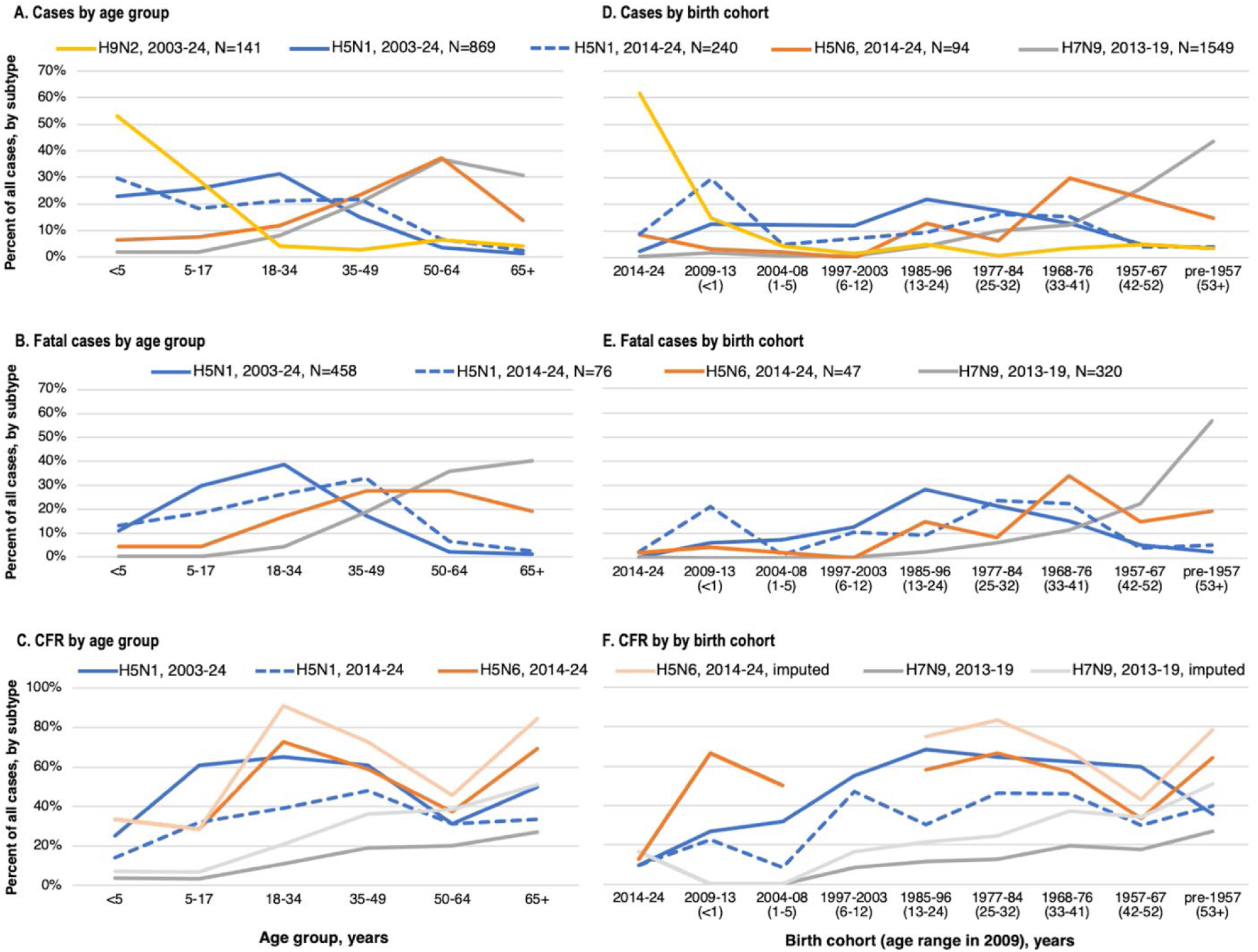
Percentage distribution of human zoonotic influenza A cases and fatalities, and case fatality ratios (CFRs) by age group and birth cohort. Displayed are the percentage distributions by age group of (**A**) cases, (**B**) fatalities and (**C**) case fatality ratios (CFR, %) of human zoonotic influenza A cases with known age including H9N2, H5N1, H5N6 and H7N9, with the same displayed instead by birth cohort in panels (**D-F).** Percentage distribution of H9N2 fatalities and CFRs not displayed owing to few such fatal cases (n=3). Legends include the span of occurrence years, with the dotted blue line indicating H5N1 distributions restricted to cases occurring since 2014. In panels D-F, values specified under the x axis provide the age range that the specified birth cohorts would have been in 2009 (during the 2009 pandemic). As lighter colored lines in panels C and F, we also display the imputed H5N6 and H7N9 CFRs assuming fatal cases with missing age follow the same distribution as fatalities for whom age and birth year were known. Absolute case and fatality tallies by age group and birth cohort are provided in **Supplementary Figure 3**.

Pre-1968 birth cohorts are substantially under-represented among H5N1 (8% and 8%) versus H7N9 (70% and 79%) cases and deaths, notably including cohorts born pre-1957 for H5N1 (4% and 2%) versus H7N9 (43% and 57%) (**Figs. 2 and 4, Supplementary Fig. 3**).

Distributions by birth year overlap between 1968-1976 when the first childhood infection may have been H3N2 or H1N1, but diverge on either side, with relative H5N1 paucity before and H7N9 paucity after (**Fig. 2**). Patterns persist with restriction to cases occurring since 2014 (**Fig. 3**) or between 2013-17 (**Supplementary Fig. 4**), but spike in post-2009 birth cohort contribution to H5N1 becomes more evident with such restriction, as does a relative notch in H5N1 versus H7N9 contribution around the 1977 H1N1 pseudo-pandemic. Birth cohorts thereafter contribute consistently more to H5N1 than H7N9, with declining trend in H5N1 contribution among birth cohorts leading up to the 2009 H1N1 pandemic (**Figs. 3 and 4, Supplementary Fig. 4**).

Fatalities show similar patterns but with more variability given reduced tallies (**Fig. 4, Supplementary Fig. 3, Supplementary Fig. 5**).

## H5N6 also older, but H9N2 younger, than H5N1 cases

Within our dataset, H5N6 cases have median (IQR) age of 50 (33-55) years and birth year of 1971 (1965-85), intermediate between H5N1 and H7N9 (**Figs. 2-4, Supplementary Fig. 3**). Conversely, H9N2 cases have median (IQR) age of 4 (2-8) years and birth year of 2016 (2010-19), much younger than H5N1 and all other zoonoses. Individuals <18 years comprise fewer cases and deaths due to H5N6 (14% and 9%) than H5N1 (>40%) but more cases due to H9N2 (82%) than either H5 subtype. Conversely, adults ≥50-years comprise more cases and deaths due to H5N6 (51% and 47%) and H9N2 (11%) (p=0.02) than H5N1 (≤5%), with ≥65-year-olds similarly comprising more H5N6 (14% and 5%) and H9N2 (4%) (p=0.04) than H5N1 (1%) cases and deaths. Age-related patterns are similar with case restriction since 2014 (**Fig. 3**) or between 2014-19 (overlapping span of H5N6 and H7N9 cases) (**Supplementary Fig. 4**), and among fatalities (**Fig. 4, Supplementary Fig. 3, Supplementary Fig. 5**).

Overall, pre-1968 birth cohorts comprise more than one-third of H5N6 cases and deaths (37% and 34%), mid-range compared to their H5N1 (<10%) or H7N9 (>70%) contribution (**Figs. 2-4, Supplementary Figs. 3**-4). With case restriction since 2014, percent contribution declines for both H5N1 and H5N6 on either side of 1968-76, but with less contribution by pre- 1957 cohorts to H5N1 (4% and 5%) than H5N6 (15% and 19%) cases (p=0.002) or deaths (p=0.002) (**Figs. 3 and 4**). With that restriction, contribution by the 2004-08 birth cohort, representing children who would have been pre-school-aged during the 2009 pandemic, is comparable for H5N1 (5% and 1%) and H5N6 (2% and 2%) cases (p>0.05) and deaths (p>0.05) but contribution by those born after the 2009 pandemic is higher for H5N1 (38% and 24%) than H5N6 (12% and 6%) cases or deaths (p=0.01). A comparably low proportion of H9N2 and H5N1 cases were born pre-1968 (9% vs. 8%, respectively) or pre-1957 (4% vs. 4%, respectively) (both p>0.05). Most H9N2 cases (77%) and 3/3 fatalities were born since 2009, far exceeding their contribution to H5N1 cases and deaths overall (15% and 7%) or with restriction since 2014 (38% and 24%) **(Figs. 2-4).**

## Only H7N9 CFRs highest for oldest adults

Only H7N9 shows steady increase in CFR with age, highest in the oldest/earliest pre- 1957 birth cohort (**Fig. 4, Supplementary Fig. 3**). For H5N1 and H5N6, CFRs are also lower among children than adults but both H5N1 and H5N6 show CFR decrease among adults born during the 1957-67 H2N2 epoch. Thereafter, among older adults ≥65 years or born pre-1957, H5N1 CFRs remain lower while H5N6 CFRs increase rather like H7N9. Between successive periods before and after the 2009 pandemic, whether including or excluding asymptomatic cases or those missing required age information, overall H5N1 CFRs fall substantially, from about two-thirds fatal in 2003-08 (63% and 66%, respectively), to half between 2009-13 (53% and 53%, respectively) (p=0.01 and p=0.001, respectively), and less than one-third between 2014-24 (25% and 32%, respectively) (**Fig. 5, Supplementary Note 1**). Among those with known age, in no period were H5N1 CFRs highest in the oldest adults (**Fig. 5**). CFR reduction in 2009-13 vs. 2003-08 was driven by adults ≥50 years (58% relative reduction) among whom there was a paucity of cases or deaths (1/6; 17%) with none aged ≥65 years. General paucity of cases ≥50- years persisted through 2014-24 but with ≥20% relative CFR reductions compared to 2003-08 affecting all age groups, including CFRs halved or more among those <5, 5-17 and ≥65 years.

**Fig. 5.**
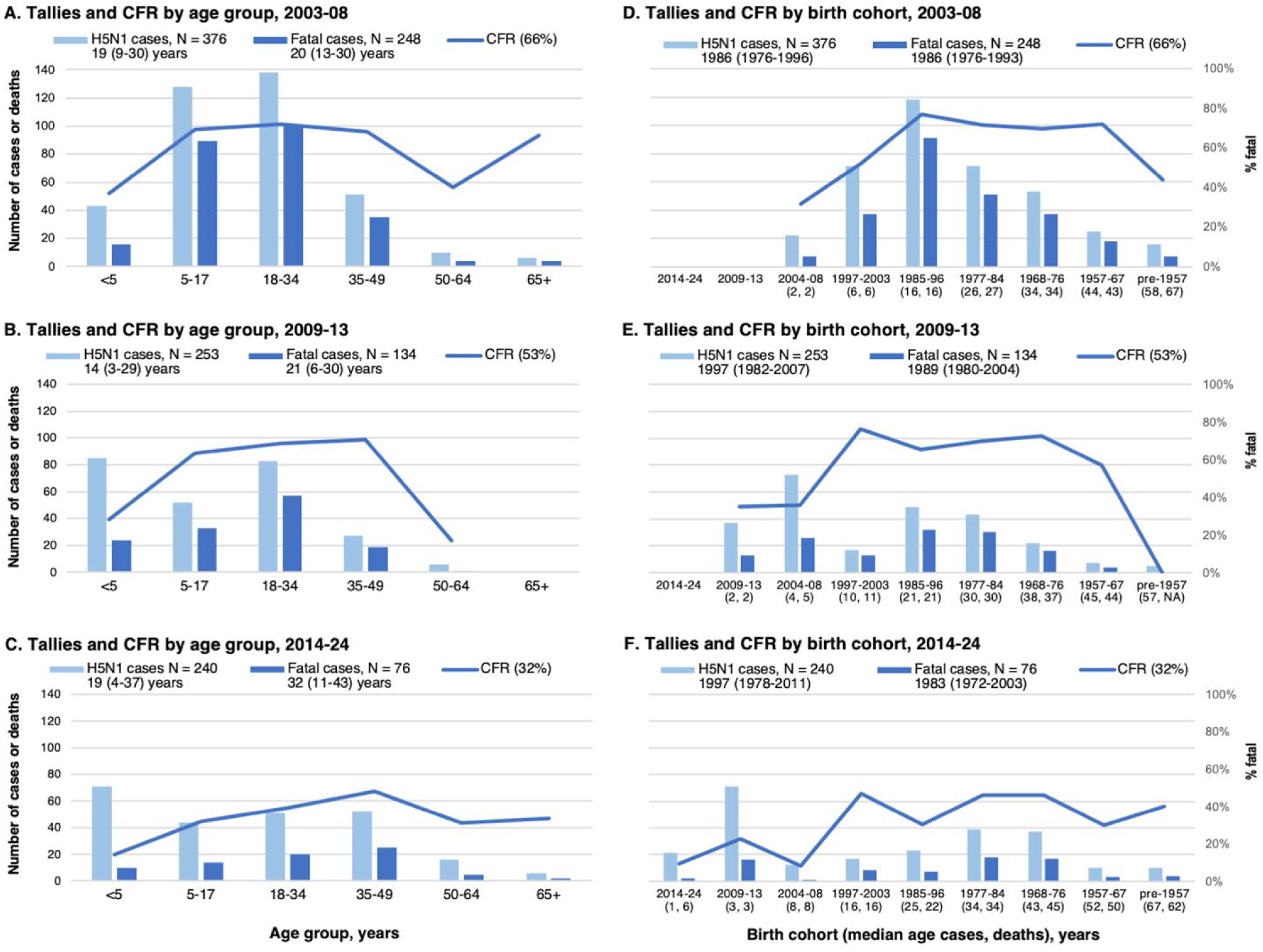
Zoonotic influenza A(H5N1) case and death tallies and case fatality ratios by age group, birth cohort and period. Displayed are tallies of laboratory-confirmed zoonotic H5N1 cases and known deaths (bars), and case fatality ratios (CFR, %) (lines), by known age group and case period (**A**) 2003-2008 (i.e., before the 2009 A(H1N1) pandemic); (**B**) 2009-2013 (during or shortly following the 2009 pandemic); and (**C**) 2014-2024 (inclusive of later seasonal H1N1pdm09 epidemics). The same information is displayed by birth cohort in panels (**D-F).** Legends include total number of cases and deaths and below that median (interquartile range) of age and birth cohort, with overall CFR specified in parentheses (among those with known age, for overall CFRs regardless of age see **Supplementary Note 1**). Numbers in parentheses below specified birth cohorts represent corresponding median ages of cases and deaths, respectively.

Within the pre-1957 birth cohort, H5N1 CFRs were comparable in 2014-24 (40%) and 2003-08 (44%) (p>0.05) despite advancing age, comparable also to younger adult birth cohorts despite overall CFR reductions.

## Discussion

In this updated review of publicly available genomic and surveillance data we report varying age-related risk by zoonotic influenza A subtype that aligns not only with previously hypothesized HA2 but also NA imprinting effects. Fundamentally underpinning our analyses is the recognition that zoonotic influenza risk in humans is directly related to the likelihood of exposure to infected animals. That we observe the greatest proportion of zoonotic cases due to H9N2 avian influenza in the very young and conversely due to H7N9 in the very old, reinforces that animal exposures occur at both extremes of age. In that context, the paucity of H5N6 and H7N9 in the very young and, conversely, of H5N1 and H9N2 in the very old, suggests some differential protective factor modulating the age-related risk. We hypothesize this protective factor to be pre-immunity induced not only by homo-group HA2 but also homosubtypic NA imprinting to subtypes variously circulating in the human population over the past century.

Highest H7N9 CFR at oldest age is consistent with prior hypotheses of predominant H3 imprinting to shared group 2 HA2 epitopes, cross-protecting against H7 among younger cohorts born since the 1968 H3 pandemic^44^. Our genetic identity analyses, however, do not spotlight exceptional H7 and H3 stalk identities, ranging 60-65%, and the same age-related increase in severe outcomes occurs with non-influenza respiratory viruses^117,120,125^, without invoking imprinting phenomena. More exceptional to explain is the pattern of decreased H5N1 risk among older adults, not only the 1957-67 birth cohort imprinted to group 1 H2N2, for whom H5N6 risk is also lower, but notably also the oldest pre-1957 birth cohort imprinted to group 1 H1N1, for whom H5N1 and H5N6 risks diverge.

We present several lines of ecological evidence to support homosubtypic anti-NA effects contributing to varying age-related zoonotic risk. Firstly, H5N1 shares at least as much, if not more, identity in the homosubtypic NA head as in the homo-group HA2 stalk with pre-1957 H1N1 viruses. Highest NA head identity, exceeding 90%, is notable in relation to the N1 of 1918 and phylogenetically-related H1N1pdm09 strains, with correspondingly lowest CFRs among the oldest cohorts and those who were pre-school or school-aged during the 2009 pandemic, the latter experiencing the highest attack rates^20^. The H1N1pdm09 pandemic would have provided original pediatric priming but also massive boost opportunity for older cohorts primed as children during more distant but phylogenetically related H1N1 epochs^18^. Whether due to HA2 and/or NA imprinting, we show overall decrease in H5N1 CFRs following the 2009 H1N1 pandemic. A greater proportion of H5N1 than H5N6 cases were born after the 2009 pandemic, suggesting less protective influence of the pandemic on H5N6 despite shared H5. Also in sharp contrast to H5N1, H5N6 CFRs increase among pre-1957 birth cohorts, a pattern more comparable to H7N9 similarly NA heterosubtypic to all human subtypes. Finally, H9N2 cases show the mildest and youngest profile of all zoonotic cases, dramatically concentrated among cohorts born post-2009 while dramatically sparing those with the greatest accumulated homo- group 1 (H1, H2) but also homosubtypic N2 (H2N2, H3N2) imprinting opportunities. Moreover, despite shared HA group 1, a greater proportion of H9N2 than H5N1 cases were born pre-1957.

Our observations are consistent with experimental evidence showing anti-N1 antibodies cross-react against H5N1, notably when induced by H1N1pdm09 strains^26,41,66,126–129^. In animal models, protection against H5N1 challenge has been attributed to anti-N1 effects also notably induced by H1N1pdm09 inoculation^59,67,126,130–135^. Similarly, anti-N2 antibodies induced by H2N2 and H3N2 pandemic strains cross-react and protect mice against H9N2 challenge^136^.

Although sero-protective anti-NA thresholds have yet to be established, recent serosurveys in Thailand^137^, Hong Kong^138^, and among US military personnel^139^, show a substantial proportion of the general population with pre-existing, cross-reactive anti-N1 to H5N1, also notably higher among sera collected after the 2009 H1N1 pandemic^138,139^. Consistent with the reduced H5N1 CFRs we observe in relation to the 2009 pandemic, anti-(H5)N1 levels are highest in the oldest cohorts, increased after confirmed H1N1pdm09 infection or vaccination, and strongly correlated with anti-H1N1pdm09 antibody. The extent to which such pre-immunity may have mitigated human H5N1 adaptation to date, or may explain host-specific variation in disease severity (including alarming mortality among presumably influenza-naïve animals)^101,102,104^, remains speculative. We acknowledge other factors contribute to infection risk and virulence such as exposure intensity, inoculum or route as well as viral genotypic (e.g., D1.1 vs. B3.13) or mutational markers^140,141^. Clade 2.3.4.4b is also distinguished from earlier H5N1 viruses by a longer NA stalk, potentially influencing host adaptation, severity, and expansion^83,142^.

We wish to emphasize that pandemic risk assessment for emerging influenza and other highly changeable viruses subject to birth (immunological) cohort effects requires granular age information, sufficient at least to derive birth year. Ideally, a central repository would maintain open-access line-lists of human zoonotic cases globally, regularly updating key variables including age and conclusive outcome resolution. Routine surveillance tends to over-estimate CFRs through disproportionate under-ascertainment of mild or asymptomatic cases, a bias paradoxically exacerbated by the infection-permissive but disease-attenuating effects of anti-NA we sought to assess. In that regard, the protective effects of anti-NA imprinting we hypothesize among particular birth cohorts may be even greater than discerned through surveillance data. To partially address this denominator-driven instability, we also explored case and death distributions separately. Conversely, CFRs based upon known deaths will be under-estimated by missing outcome information. Our crude analyses do not account for varying exposure opportunities overall, by age, place or time, and we did not derive incidences, given further uncertainty in estimated populations at risk, including cumulatively over time. Overall, we emphasize patterns over absolute estimates, with our H5N1 and H7N9 patterns regardless comparable to earlier Gostic et al modelling^44^. Linear percent amino acid identities are a measure of relatedness that do not directly translate antigenically or immunologically. Finally, imprinting epochs based on birth year will result in some misclassification given variable (5-10 year) delays to the age of first influenza infection^10,11,35,55^, a consideration most relevant to those born just prior to epoch transition.

Better understanding of pre-immunity against emerging avian influenza has become more critical in the context of the now more ubiquitous H5N1 threat. Whereas conventional serological assays target the HA (e.g., hemagglutination inhibition), our findings reinforce the need for greater access and deployment of anti-NA assays to better quantify and compare homo- and heterosubtypic NA responses^143^. Homosubtypic anti-NA in particular may have attenuating effects on H5N1 and H9N2 zoonotic risk, notably severity, with implications for targeted messaging and mitigation measures. Alternatively, while beneficial to those directly exposed, anti-NA may also paradoxically contribute to unrecognized infections, potentially facilitating surreptitious shedding, spreading and viral adaptation, with implications for case detection, containment and enhanced surveillance. To capitalize on broader facets of pre-immunity including back-boost of protective imprints to several viral domains, the preferred human zoonotic influenza vaccine might include antigen closely matched to the target HA1, while also HA2 homo-group and NA homosubtypic to human adapted subtypes. As practical example, all things being equal, H5N1 may be preferred to H5N8 candidate vaccine options for pre-pandemic deployment against H5N1^144^. To date, however, both seasonal and pandemic vaccines are only standardized for HA antigen content^145^. In the context of universal seasonal influenza immunization programs and their potential interaction with imprinted pre-immunity, our findings reinforce additional standardization of NA content and quantification of their effects in seasonal and pandemic vaccines^26–28,146^. Ultimately, in the event of a pandemic, advance understanding of the variability in pre-existing immunity will be important to guide prioritization of scarce supplies such as vaccines or antivirals. Overall, our review based upon updated genetic and surveillance data show age and birth cohort distributions differ dramatically by subtype of human zoonotic influenza, aligning not only with homo-group HA2 but also homosubtypic NA imprinting epochs and effects. Given the critical importance to pandemic risk assessment and response, and the escalating threat posed by H5N1, the cross-protective role of both HA and NA imprinting against emerging influenza zoonoses warrants urgent and definitive investigation.

## Data Availability

All analyses are based upon publicly available data with references and links provided in supplementary information. Those seeking additional details may also contact the corresponding author.

## Methods

### Genetic relatedness analyses

The genetic relatedness analyses earlier presented by Gostic et al^44^ were predicated on the percentage of amino acids *similar* in pairwise comparison of the hemagglutinin (HA) stalk (HA2) segment of H5N1 and H7N9 avian influenza versus human influenza A subtypes. They cited high HA2 similarity for avian H5 with group 1 H1 (94%) and H2 (98%) human subtypes, and for avian H7 with the group 2 H3 (86%) human subtype. Our analyses are instead predicated upon the percentage of amino acids that are *identical* in pairwise comparison of HA2 but also HA1 head and neuraminidase (NA) head domains. *Identity* analyses are simpler, more specific and less susceptible to peptide *similarity* definitions, while also more directly replicable and reliable to interpret, especially at higher percentages; however, they may be less sensitive to functional or biochemical relatedness at lower percentages^147^.

We derived percent pairwise identities in matched HA1, extracellular HA2 and NA head domains of specified influenza A viruses as the number of identical amino acid residues between paired sequences divided by the total number of aligned amino acid positions, excluding matching gap characters. We retrieved protein sequences from the Global Initiative on Sharing All Influenza Data (GISAID), on June 11, 2024 spanning sample collection dates from January 1, 1918 to February 29, 2024^124^. After removing duplicate sequences, and those with incomplete coverage or of swine origin, we conducted pairwise analysis of avian and human zoonotic strains in relation to human adapted (pandemic or seasonal) strains, including: (a) 93,194/125,879 HA and 85,774/108,033 NA sequences from human H1N1 (1918-2024); (b) 44/50 NA sequences from human H2N2 (1957-1968); and (c) 114,006/169,064 HA and 101,684/135,740 NA sequences from human H3N2 (1968-2024)^148^. Sequences with outlier identities compared to yearly medians were blasted against the NBCI blastp database and excluded if related to temporally-distant strains or products of molecular cloning^149^.

We plot individual identities in relation to human strains by year of collection of the human strain, with epoch-defining and/or major antigenic transition years for human strains noted on x-axes of **Supplementary Fig. 110**–13^,150^. We fit generalized additive models by human zoonotic subtype separately before and after the 1957–77 break in human H1N1 circulation, as applicable. Owing to few sequences, we did not plot identities in relation to human H2 and include both human H2N2 and H3N2 sequences in the same N2 models since 1957. Select influenza A strains were assessed in matrix cross-comparisons in **Supplementary Table 1** (HA1), **Supplementary Table 2** (HA2) and **Supplementary Table 3** (NA), with their GISAID identifiers provided in **Supplementary Table 5**^124^.

### Epidemiological analyses

We estimated birth year of individual human zoonotic cases by subtracting known age in years from occurrence year, the latter based hierarchically on year of onset or else severe outcome or else first official case report date. Year of fatal cases was based on case year. We excluded cases missing exact age in years. We categorized birth cohorts in relation to human subtype circulation and potential HA2 and/or NA imprinting epochs, anchoring as per Gostic et et al^44^ foremost upon the pre-1957 H1N1, 1957-1967 H2N2 and post-1968 H3N2 periods of subtype predominance.

We subdivided the post-1968 H3N2 epoch for more comparable time spans with additional rational for sub-categories as follows: (1) 1968-76 (sole H3N2 circulation); (2) 1977- 84 (H1N1 pseudo-pandemic due to “A prime” like virus and co-circulation thereafter of descendant H1N1 and H3N2 strains)^10–13^; (3) 1985-96 (ongoing co-circulation of “A prime” H1N1 and H3N2 descendant strains) ^10–13^; (4) 1997-2003 (H3N2 epidemics spanning A/Sydney/5/97 through A/Fujian/411/2002 major antigenic cluster transitions and global circulation of human-origin H1N2 reassortment virus 2000-2003)^14,150,151^; (5) 2004-08 (continued H1N1 “A prime” and H3N2 descendant virus co-circulation preceding 2009 H1N1pdm09 pandemic); (6) 2009-13 (H1N1pdm09 pandemic, virus related to the more distant “Asw” era, co-circulation with H3N2; 2013-14 seasonal epidemic due to antigenically drifted H1N1pdm09 virus)^53,54^; and (7) 2014-24 (span of H5N6 human cases, additional H1N1pdm09 seasonal epidemics (2015-16, 2019-20, 2023-24, 2024-25), else H3N2 dominant seasons, recognizing pause in influenza and other respiratory virus circulation during the COVID-19 pandemic spanning 2020-22)^53,54,152^.

We plot crude percentage distributions of human zoonotic cases by age in single years or age group and by birth year or cohort. Statistical comparisons are by chi-square, Fisher exact or Wilcoxon rank-sum test as indicated. We derived case fatality ratios (CFRs) as the tally of known fatalities divided by the corresponding tally of confirmed cases, expressed as percentages, overall and by age group or birth cohort. If not definitively recovered or fatal, the former was assumed. Given a greater proportion of H7N9 and H5N6 deaths reported in aggregate for whom individual-level age could not be reconciled, we also explored their age-specific CFRs assuming fatal cases missing age follow the same distribution as fatalities with known age and birth year.

All H5N6 (94/94) and virtually all H9N2 (135/141; 96%) and H7N9 (1394/1549; 90%) cases but fewer H5N1 (240/869; 28%) occurred since 2014 (**Fig. 1**). We thus also compared distributions with restriction to cases since 2014, between 2013-17 (>99% of H7N9 cases) and 2014-19 (first H5N6 through last H7N9 case). As evident in our assembled databases, more H5N1 cases were hospitalized between 2003-23 (734/883; 83%) than in 2024 (15/81; 19%).

However, of the 81 H5N1 cases reported in 2024, 11/13 (85%) in Asia and 4/5 (80%) identified through routine surveillance elsewhere were hospitalized; conversely, among 63 commercial animal workers in the US, none was hospitalized. To address the systematic differences in severe outcome ascertainment based upon enhanced occupational vs. routine severe outcome surveillance, we excluded cases known to be asymptomatic and/or commercial animal workers.

## Acknowledgments

Authors acknowledge the patients and clinical, laboratory, public health, academic, animal health and other sources of case details assembled and analyzed in the current work. Detailed source information for each of the human zoonotic cases, by subtype, is provided in Supplementary Notes 1-4. Authors also gratefully acknowledge the authors, originating laboratories, and submitting laboratories of the reference virus strains for sharing those via the GISAID Initiative, which we rely on for our sequence relatedness analyses. We thank Yuping Zhan of the BC Centre for Disease Control for assistance in verifying some case details.

## Funding

No external funding was provided for this work.

## Competing interests

DMS is Principal Investigator on grants received to her institution from the Public Health Agency of Canada in support of this work. She has received grants from Pacific Public Health Foundation and Canadian Institutes of Health Research and the Michael Smith Foundation for Health Research for unrelated work, also paid to her institution. Other authors declare that they have no competing interests.

## Data and materials availability

All analyses are based upon publicly available data with references provided in supplementary information. Those seeking additional details may also contact the corresponding author.

## Supplementary Information

Supplementary Notes 1-4

Supplementary Figures 1-5

Supplementary Tables 1-5

Supplementary Information, References

## Notes

### Author Declarations

The study used only openly available human data. All analyses are based upon publicly available data with references and links provided in supplementary information.

