## Supplementary Information for "Neuraminidase imprinting and the age-related risk of zoonotic influenza"

**Supplementary Notes [hyperlinked]**

**Supplementary Note 1:** Completeness of H5N1 dataset
**Supplementary Note 2:** Completeness of H7N9 dataset
**Supplementary Note 3:** Completeness of H5N6 dataset
**Supplementary Note 4:** Completeness of H9N2 dataset

**Supplementary Figures [hyperlinked]**

**Supplementary Figure 1:** Percent amino acid pairwise identities between select avian and human zoonotic relative to human adapted influenza A viruses, 1918-2024
**Supplementary Figure 2:** Distribution of human zoonotic influenza A H5N1 cases by state in the United States, 2022 and 2024
**Supplementary Figure 3:** Human zoonotic influenza A case and fatality tallies and case fatality ratios by age group and birth cohort
**Supplementary Figure 4:** Percentage distribution of human zoonotic influenza A cases by single year of age and birth, restricted to 2013-2017 and 2014-2019
**Supplementary Figure 5:** Percentage distribution of fatal human zoonotic influenza A cases by year of age and birth year, overall and since 2014

**Supplementary Tables [hyperlinked]**

**Supplementary Table 1:** Hemagglutinin (HA) head (HA1): percent amino acid pairwise identities between select influenza A reference viruses
**Supplementary Table 2:** Hemagglutinin stalk (HA2): percent amino acid pairwise identities between select influenza A reference viruses
**Supplementary Table 3:** Neuraminidase (NA) head: percent amino acid pairwise identities between select influenza A reference viruses
**Supplementary Table 4:** Global distribution of human zoonotic H5N1 cases by country, included in line-list overall and by period, 1997-2024
**Supplementary Table 5:** Neuraminidase (NA) and hemagglutinin (HA) sequences of reference viruses used in pairwise identity comparisons obtained from the Global Initiative on Sharing All Influenza Data (GISAID)

**Supplementary Information, References [hyperlinked]**

### **Supplementary Note 1:** Completeness of H5N1 dataset

Absent an officially posted, publicly accessible line-list of human H5N1 case details, we assembled our own individual-level line-listed H5N1 database. We drew from various public sources including the World Health Organization (WHO) cumulative tallies (as per January 20, 2025, report through December 31, 2024)[1], WHO risk assessments and summaries of influenza at the human-animal interface (monthly)[2], WHO disease outbreak news[3], WHO timeline of major events in animals and humans, 1996 to 2014[4], Western Pacific Region (WPR) weekly reports (as of February 20, 2025, number 986)[5] and other sources such as Center for Infectious Disease Research and Policy (CIDRAP)[6], ProMED[7], FluTrackers[8], peer-reviewed literature and media. We particularly acknowledge Fiebig et al, 2011[9] for their line-list of confirmed cases between September 2006 and August 2010. As per WHO, we only include cases from 2003 and our dataset spans through December 31, 2024. We reconciled completeness of our H5N1 database, notably for age and fatal status as follows:

| **Period** | **H5N1 case and fatality counts by period and source, including line-list tallies overall and with exact age in years** | | | | | | | | |
|---|---|---|---|---|---|---|---|---|---|
| | WHO cumulative tallies (2003 – December 31, 2024)[*] | | | Line-list (as of December 31, 2024) | | | Line-list, with exact age in years (as of December 31, 2024) | | |
| | **Cases** | **Deaths** | **CFR %** | **Cases** | **Deaths** | **CFR %** | **Cases** | **Deaths** | **CFR %** |
| 2003-2008[†,‡] | 395 | 250 | 63% | 395 | 248 | 63% | 376 | 248 | 66% |
| 2009-2013[§,**] | 254 | 135 | 53% | 255 | 135 | 53% | 253 | 134 | 53% |
| 2014-2019[††] | 212 | 70 | 33% | 212 | 67 | 32% | 212 | 67 | 32% |
| 2020-2023[‡‡] | 21 | 6 | 29% | 21 | 6 | 29% | 16[§§] | 6 | 38% |
| 2024 | 81 | 4 | 5% | 81 | 4 | 5% | 15[***] | 3 | 14% |
| **2003-2023** | 882 | 461 | 52% | 883 | 456 | 52% | 857 | 455 | 53% |
| **2003-2024** | 963 | 465 | 48% | 964 | 460 | 48% | 872 | 458 | 53% |
| **2014-2024** | 314 | 80 | 25% | 314 | 77 | 25% | 243 | 76 | 31% |

Earliest case in our H5N1 dataset spanning through December 31, 2024 and with known age and onset is a 5-year-old from Vietnam in December 2003[12], the latest is a 13-year-old from Canada in November 2024[13]. Exact age in years, sex and onset information were not available for any of the 67 cases (including one in 2022) reported by the United States through December 23, 2024. Our analyses are otherwise complete for exact age in years for: 872/963 (91%) cases and 458/465 (98%) known fatalities between 2003-2024 and for 243/314 (77%) cases and 76/80 (95%) known fatalities between 2014-2024. Additionally excluding asymptomatic cases identified through enhanced screening/surveillance, we include 868 cases and 458 deaths in single year of age/birth analyses between 2003-2024 and 239 cases and 76 deaths between 2014-2024. In grouped age or birth year categories we add a 2024 US daycare attendee <5 years (born 2020-2024), thus including 869 cases in total and 240 between 2014-2024.

---

[*] As per WHO cumulative tallies reported January 20,2025 through December 31, 2024. Note that as per WHO, the 18 cases (6 deaths) in 1997 are excluded but case details are available from Lai et al, 2016[10], and Chan, 2002[11]

[†] 2 deaths missing within our line-list, both among cases in Vietnam in 2005 that could not be assigned.

[‡] 19 missing age information within our line-list: 17 from Vietnam in 2005; 2 from Pakistan in 2007.

[§] Our line-list includes one additional case from Egypt in 2012 compared to WHO (same number of fatalities for Egypt in 2012).

[**] 2 missing age information within our line-list, both from Egypt in 2011 including 1 specified as “young child” and 1 as “adult.

[††] 3 deaths missing within our line-list, all among cases in Egypt in 2015 that could not be assigned.

[‡‡] 5 missing age information within our line-list, 4 from the United Kingdom (UK) in 2023 specified as adult worker; 1 from the United States (US) in 2022 specified as 19-39-year-old.

[§§] Among these 16 cases with known age in years are 4 asymptomatic infections identified through enhanced screening. They are excluded from age and birth year analyses. Thus, 12 included in single year analyses for 2020-2023, 27 for 2020-2024, 853 for 2003-2023, 868 for 2003-2024 and 239 for 2014-2024.

[***] Of the 66 cases in 2024 missing exact age in years, all (including one fatality) are from the US with report dates through December 23, 2024. In age group/birth cohort analyses we additionally include one non-fatal US case with symptom onset on November 11, 2024 reported from California as a pediatric daycare attendee without known animal exposure (categorized as <5 years born 2020 to 2024). Thus, 16 included in age group/birth cohort analyses for 2024, 28 for 2020-2024, 869 for 2003-2024 and 240 for 2014-2024.

**Supplementary Note 2:** Completeness of H7N9 dataset

Absent an officially posted, publicly accessible line-list of human H7N9 case details, we assembled our own individual-level, line-listed H7N9 database. We drew from various public sources including the World Health Organization (WHO) WHO disease outbreak news[3], Western Pacific Region (WPR) weekly reports (as of February 20, 2025, number 986)[5], the Hong Kong Department of Health weekly avian influenza reports (as of February 22, 2025, volume 21, number 8)[14], and FluTrackers[8]. As per WHO, we only include cases from 2013 and our dataset spans through December 31, 2024, although the last reported H7N9 case was in April 2019. We reconciled completeness of our H7N9 database, notably for age and fatal status as per below.

Since early 2013, the Western Pacific Region (WPR) weekly reports (as of February 20, 2025, number 986)[5] indicate 1,568 laboratory-confirmed human infections with avian influenza H7N9 virus reported to the WHO, with the last reported case having onset in 2019. Overall, 616/1568 (39%) are reported by the WHO to have been fatal; however, a substantial number of deaths were reported in aggregate only.

As noted by others, our line-list shows the majority (1537/1568; 98%) of cases reported from mainland China, with others imported from mainland China as follows: Hong Kong (21), Taiwan (5), Canada (2), Macao (2), Malaysia (1)[14,15].

Earliest case in our H7N9 dataset with known age and onset is an 87-year-old from China in February 2013[16], the latest is an 82-year-old from China in March 2019[17].

Our line-list is complete for exact age in years for 1549/1568 (99%) H7N9 cases and includes 324/616 (53%) known fatalities of which 320/616 (52%) fatalities have known exact age in years.

### **Supplementary Note 3:** Completeness of H5N6 dataset

Absent an officially posted, publicly accessible line-list of human H5N6 case details, we assembled our own individual-level line-listed H5N6 database. We drew from various public sources including the World Health Organization (WHO) disease outbreak news[3], Western Pacific Region (WPR) weekly reports (as of February 20, 2025, number 986)[5] and Center for Infectious Disease Research and Policy (CIDRAP)[6], ProMED[7], FluTrackers[8], peer-reviewed literature and media. As per WHO, we only include cases from 2014 and our dataset spans through December 31, 2024. We reconciled completeness of our H5N6 database, notably for age and fatal status as per below.

**2014-2021: 67 cases, 37 deaths**

In aggregate, Zhu et al, 2022[18] describe 65 human H5N6 cases with onset between April 21, 2014 and December 31, 2021, as per China's national surveillance system. We verified age, onset year, geographic and known fatal status in relation to the 65 aggregate cases they described during this period. However, our line-list includes 67 H5N6 cases with onset between 2014 and 2021, with the following discrepancies:

1. Zhu et al, 2022[18] do not include our case 01. This 5-year-old from China with onset in February 2014 (recovered) was reported by Zhang et al, 2016[19] and retrospectively by the WHO summary and risk assessment (as of 20 January 2016)[2].
2. Zhu et al, 2022[18] include our case 06. This 4-year-old from China with onset in October 2015 (fatal) was reported by Li, T et al, 2016[20] but is not officially included in WHO tallies.
3. Zhu et al, 2022[18] do not include our case 32. This 5-year-old from Lao People's Democratic Republic with onset in February 2021 (recovered) was confirmed by the WHO summary and assessment (from 30 January to 15 April 2021)[2].

Zhu et al, 2022[18] report 36 deaths among 65 cases including 18 deaths before 2021 and 18 in 2021. We individually linked all36 deaths, including 18/18 before 2021 and 18/18 in 2021. Based on the subsequently published systematic review of H5N6 cases (Li, F et al, 2024)[21] and verification against original case report (by Meng et al, 2018)[22], we identified one additional death in a 30-year-old in 2016, increasing the total to 37 identified deaths among 67 cases from 2014-2021.

**2022-2024: 27 cases, 10 deaths**

For the period spanning 2022 to 2024, we assessed completeness in relation to individually reported cases from Western Pacific Region (WPR) weekly reports (as of February 20, 2025, number 986)[5]. With last onset date of June 17, 2024 involving a 70-year-old from Anhui, China[23], our case tally (n=27) aligns with the WHO. Our death tally (n=10) exceeds the WHO (n=7) for this period, owing to retrospective reporting of deaths in published literature from China by Li, L et al, 2024[24] (54-year-old from Hunan), Li, F et al, 2024[21] (28-year-old from Henan), both (verified against original case report by Jia et al, 2023[25]), as well as Lin S et al, 2024[26] (51-year-old from Jiangxi, China) all in 2022. Age information is complete for all cases and deaths.

**2014-2024 completeness as of December 31, 2024: 94/94 cases; 47/57 deaths**

WHO official tallies specify 93 laboratory-confirmed cases as per Western Pacific Region (WPR) weekly reports (as of February 20, 2025, number 986)[5]; whereas, we include 94 H5N6 cases. All but the case from Lao PDR in 2021 were reported from China. All cases in our dataset spanning through December 31, 2024 are complete for age, beginning with the 5-year-old with onset in February 2014[19], through the 70-year-old with onset in June 2024[23]. The WHO WPR Weekly Updates between July 19 (number 956) and July 26, 2024 (number 957) indicate 38 individually tracked deaths[5]; however, in the latter report, the WHO WPR also retrospectively added 19 deaths without age information, tallying 57 deaths in total (i.e., 57/93; 61%). We captured a total of 47 H5N6 fatalities each with known age in years. Our line-list is thus complete for age for 44/57 (82%) deaths as of December 31, 2024.

### **Supplementary Note 4:** Completeness of H9N2 dataset

Absent an officially posted, publicly accessible line-list of human H9N2 case details, we assembled our own individual-level line-listed H9N2 database. We drew from various public sources including the World Health Organization (WHO) disease outbreak news[3], WHO risk assessment and summary of influenza at the human-animal interface (monthly)[2], FluTrackers[8], and Western Pacific Region (WPR) weekly reports (as of February 20, 2025, number 986)[5] as well as the Hong Kong Department of Health weekly avian influenza reports (as of February 22, 2025, volume 21, number 8)[14] and GISAID records[27]. As per WHO, we only include cases from 2003 and our dataset spans through December 31, 2024. We reconciled completeness of our H9N2 database, notably for age and fatal status as per below.

In WHO risk assessment and summary of influenza at the human-animal interface (13 December to 20 January 2025)[2], WHO cites “nearly 130 human infections with A(H9N2) cases have been reported to date since 2003, six of these have been severe or fatal and three of these were known to have underlying medical conditions”. Restricting cases to confirmed human H9N2 infections with WHO report dates from December 10, 2003, our line-list includes 134 confirmed H9N2 infections with WHO report dates by December 13, 2024. Inclusive of the additional data sources above, however, we tally 142 confirmed cases with onset dates from December 2003 through December 31, 2024.

As additional check on the accuracy and completeness of our H9N2 dataset, WPR weekly reports (as of February 20, 2025, number 986)[5] indicates: “Since December 2015, a total of 117 cases of human infection with avian influenza A(H9N2), including two deaths (both with underlying conditions), have been reported to WHO in the Western Pacific Region. Of these, 114 were reported from China, two were reported from Cambodia, and one was reported from Viet Nam.” Restricting our dataset in this way based upon *report by the WHO since but not including* December 2015 through February 20, 2025, we similarly tally 117 H9N2 cases, including 114 from China, two from Cambodia and one from Vietnam (each complete for exact age in years). Our line-list captures additional cases outside the WPR including Bangladesh (1), Egypt (1), Ghana (1), India (2), Oman (1), Senegal (1).

We reconciled completeness of our H9N2 database, notably for exact age in years as per below.

| Country | 2003 to 2024 | | 2003 to 2008 | | 2009 to 2013 | | 2014 to 2019 | | 2020 to 2023 | | 2024 | |
|---|---|---|---|---|---|---|---|---|---|---|---|---|
| | N all cases | n exact age in years | N all cases | n exact age in years | N all cases | n exact age in years | N all cases | n exact age in years | N all cases | n exact age in years | N all cases | n exact age in years |
| Bangladesh | 3 | 3 | 0 | 0 | 1 | 1 | 2 | 2 | 0 | 0 | 0 | 0 |
| Cambodia | 2 | 2 | 0 | 0 | 0 | 0 | 0 | 0 | 2 | 2 | 0 | 0 |
| China* | 126 | 125 | 2 | 2 | 3 | 3 | 35 | 34 | 65 | 65 | 22 | 22 |
| Egypt | 4 | 4 | 0 | 0 | 0 | 0 | 4 | 4 | 0 | 0 | 0 | 0 |
| Ghana | 1 | 1 | 0 | 0 | 0 | 0 | 0 | 0 | 0 | 0 | 1 | 1 |
| India | 2 | 2 | 0 | 0 | 0 | 0 | 1 | 1 | 0 | 0 | 1 | 1 |
| Oman | 1 | 1 | 0 | 0 | 0 | 0 | 1 | 1 | 0 | 0 | 0 | 0 |
| Senegal | 1 | 1 | 0 | 0 | 0 | 0 | 1 | 1 | 0 | 0 | 0 | 0 |
| Vietnam | 1 | 1 | 0 | 0 | 0 | 0 | 0 | 0 | 0 | 0 | 1 | 1 |
| TOTAL | 142 | 141 | 2 | 2 | 4 | 4 | 44 | 43 | 67 | 67 | 25 | 25 |

* Includes Hong Kong/Hong Kong ex China.

Between 2003 and December 31, 2024, the earliest recorded onset within our dataset is a 5-year-old from Hong Kong in November 2003[28], and the latest is a 2-year-old from Sichuan, China on December 27, 2024[29]. Our analyses include 142 H9N2 cases and 3 (2%) deaths, the latter including a 57-year-old[8], and 39-year-old from China[2], as well as a 37-year-old from Vietnam in March 2024, the latter reported by Duong et al, 2025[30], but not the WHO. Of the 142 H9N2 human cases in our line list, we retrieved exact age in years for 141/142 (99%) cases and all (3/3) known deaths; for the period spanning 2014 to 2024, we include 136 cases for whom exact age in years is complete for 135/136 (99%), including the three deaths.

**Supplementary Figure 1.** Percent amino acid pairwise identities between select avian and human zoonotic relative to human adapted influenza A viruses, 1918-2024.

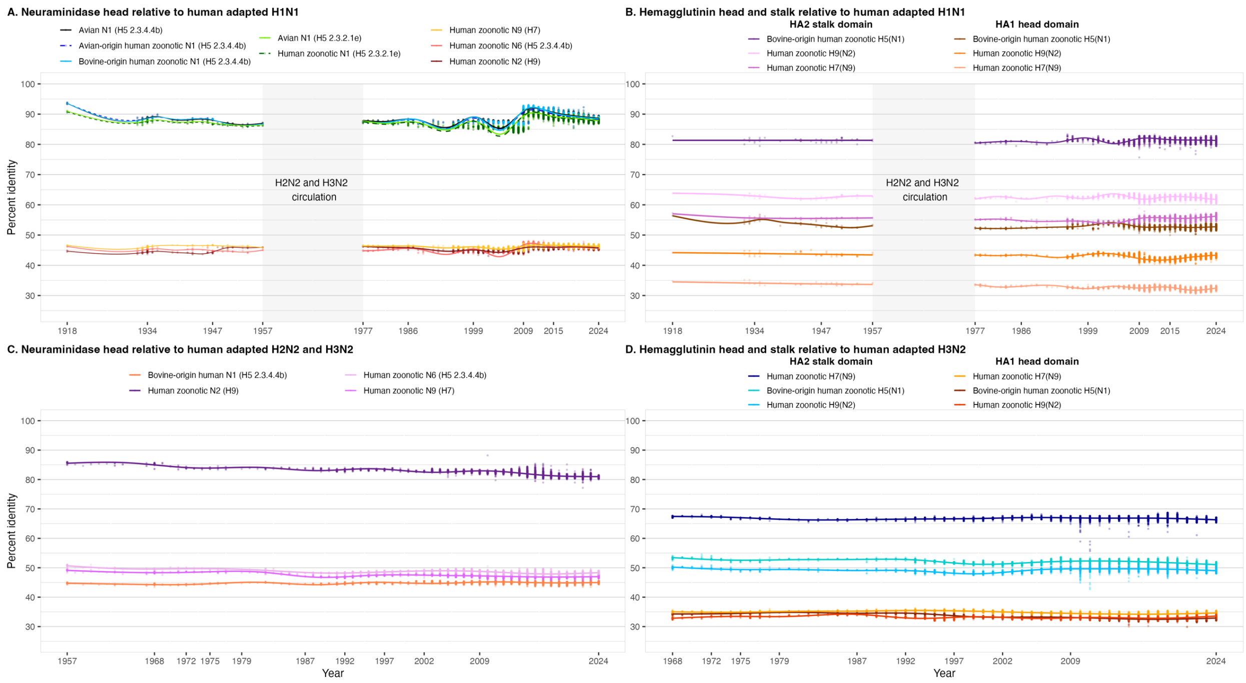


Shown by year of collection of human adapted influenza A strains on the x axis, we compare percent amino acid pairwise identities between: **(A)** neuraminidase (NA) head of human H1N1 versus representative avian and human zoonotic influenza A strains including: A/Chicken/Czech_Republic/3744-5/2024 avian N1(H5 2.3.4.4b); A/England/232780677/2023 avian-origin human zoonotic N1(H5 2.3.4.4b); A/Texas/37/2024, bovine-origin human zoonotic N1(H5 2.3.4.4b); A/Duck/Cambodia/W49h3K241D3T/2023 avian N1(H5 2.3.2.1e); A/Vietnam/KhanhhoaRV1-005/2024 avian-origin human zoonotic N1(H5 2.3.2.1e); A/Hong Kong/VM24002346/2024 avian-origin human zoonotic N2 (H9); A/Changsha/1/2022 avian-origin human zoonotic N6(H5 2.3.4.4b) and Gansu/23276/2019 avian-origin human zoonotic N9(H7); **(B)** the hemagglutinin (HA), including HA2 stalk and HA1 head of human H1N1 versus bovine-origin human zoonotic H5N1 and avian-origin human zoonotic H9N2 and H7N9 strains as per panel (A); **(C)** the NA head of human H2N2 and H3N2 versus bovine-origin human zoonotic H5N1, and avian-origin human zoonotic H5N6, H9N2 and H7N9 strains as per panel (A); and **(D)** the HA2 stalk and HA1 head of human H3N2 versus bovine-origin human zoonotic H5N1 and avian-origin human zoonotic H9N2 and H7N9 as per panel (A). Percent identities are plotted using a generalized-additive model fit for each reference virus and domain. In panels A and B, grey shading indicates absence of human H1N1 circulation, with separate regression models fit either side. Epoch-defining and/or major antigenic transition years are noted on respective x-axes. HA comparisons with human-adapted H2 are not displayed owing to few sequences.

**Supplementary Figure 2.** Distribution of human zoonotic influenza A H5N1 cases by state in the United States, 2022 and 2024.

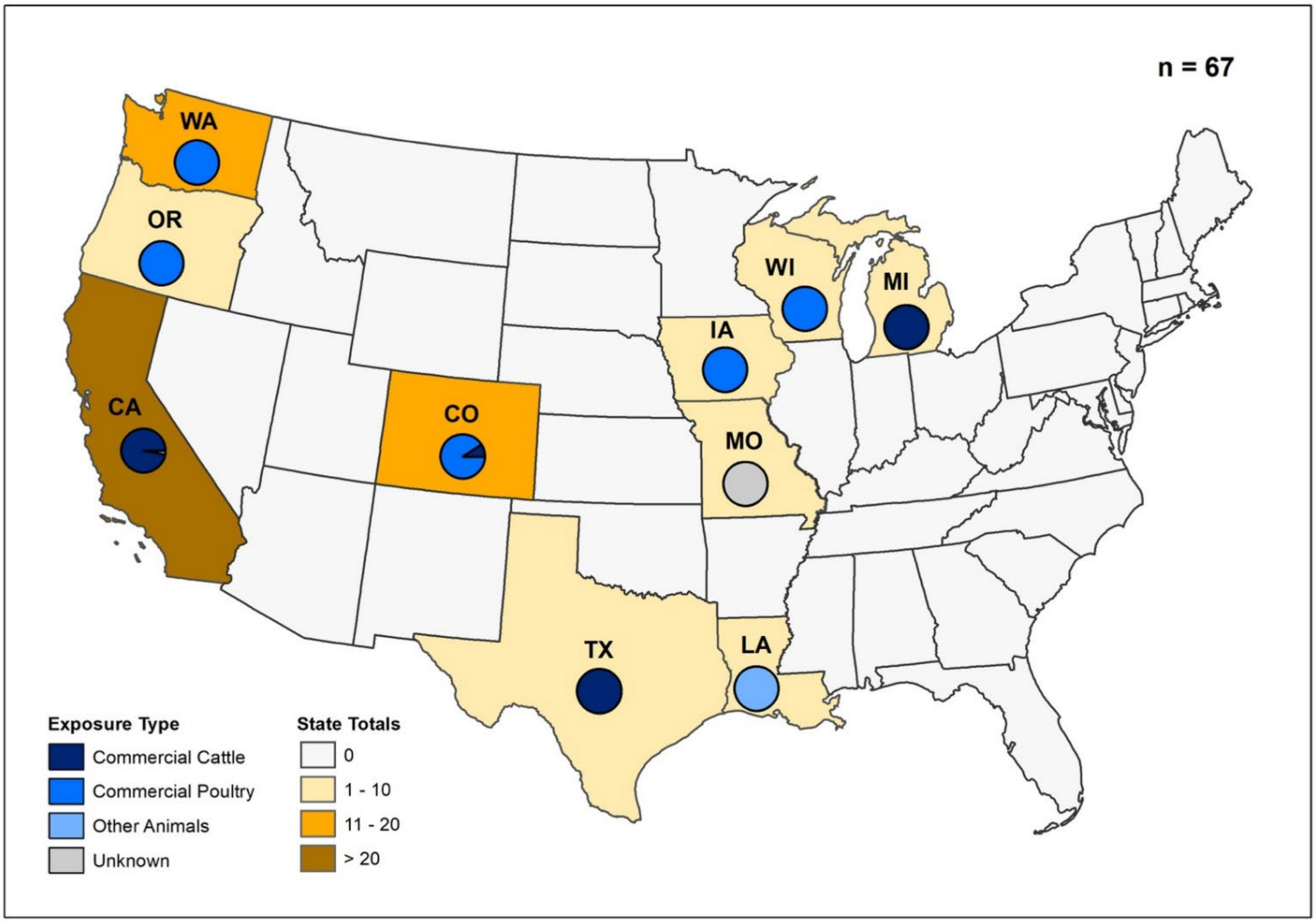


Displayed are the color-coded tallies of laboratory-confirmed human zoonotic H5N1 cases and proportionate animal source contribution (commercial cattle, commercial poultry, other animals, unknown) by state in the United States through December 31, 2024. Tallies include one case in 2022 (Texas) and 66 that occurred in 2024. Other animals may include backyard flocks, wild birds, or other mammals. See Supplementary Note 1 for further human zoonotic H5N1 case reconciliation details.

**Supplementary Figure 3.** Human zoonotic influenza A case and fatality tallies and case fatality ratios by age group and birth cohort.

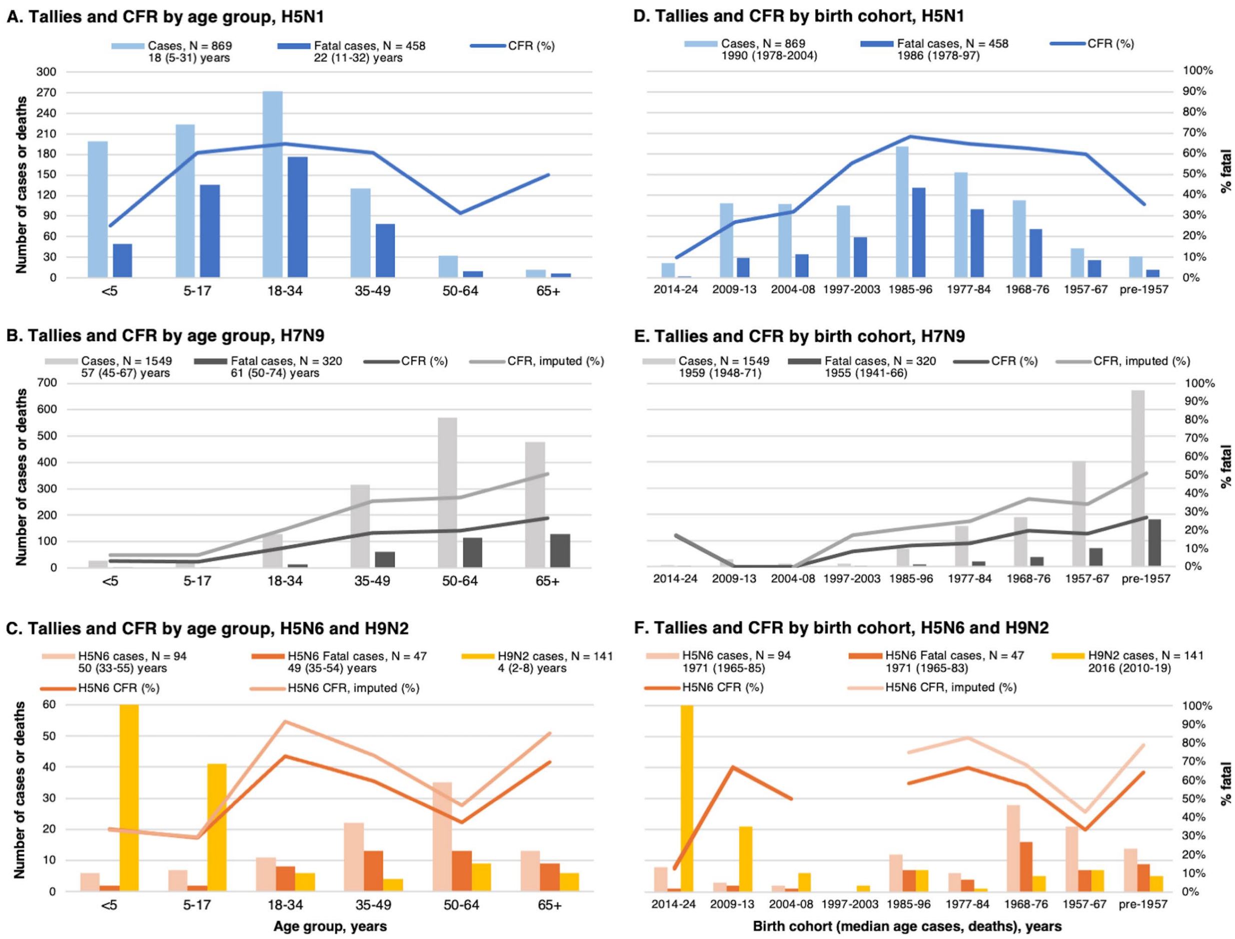


Displayed are the tallies of laboratory-confirmed human zoonotic influenza A cases and fatalities with known age through December 31, 2024, including case fatality ratios (CFR, %) by age group for (**A**) H5N1; (**B**) H7N9 and (**C**) H5N6 and H9N2 with the same displayed instead by specified birth cohort in panels (**D-F**). Legends include total number of cases and fatalities, with median (interquartile range) of age (A-C) and birth year (D-F) specified below tallies. We also display as lighter colored lines the imputed CFRs of H7N9 (panels B and E) and H5N6 (panels C and F) assuming fatal cases with missing age follow the same distribution as fatalities for whom age and birth year were known.

**Supplementary Figure 4.** Percentage distribution of human zoonotic influenza A cases by single year of age and birth, restricted to 2013-2017 and 2014-2019.

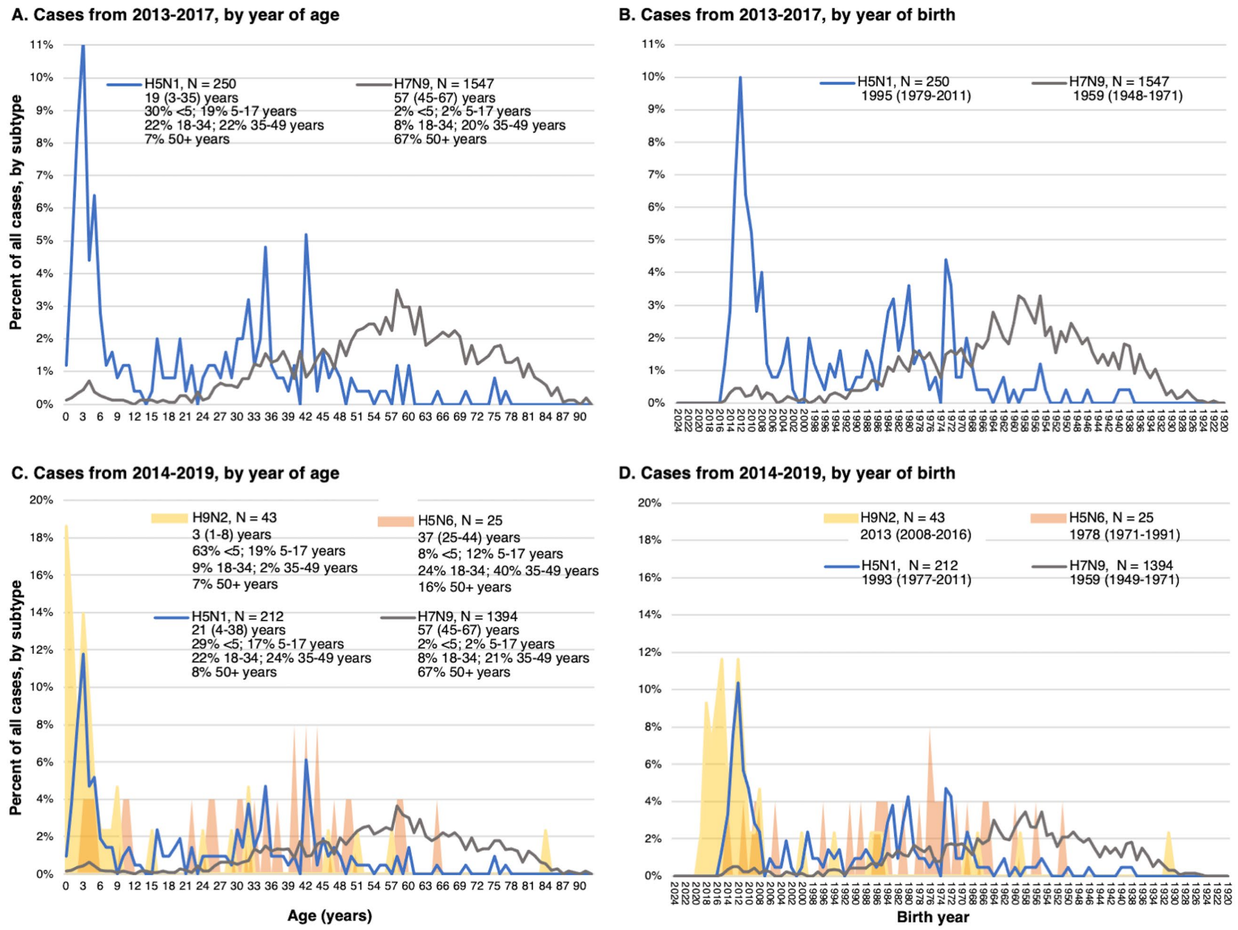


Displayed are the percentage distributions of laboratory-confirmed human zoonotic influenza A H5N1 and H7N9 cases with known age *restricted to cases between 2013-2017* (span of most H7N9 cases) by (**A**) year of age and (**B**) birth year. Percentage distributions of laboratory-confirmed human zoonotic influenza A H9N2, H5N1, H5N6 and H7N9 cases with known age *restricted to cases between 2014-2019* (overlapping span of H5N6 and H7N9 cases) are also displayed by (**C**) year of age and (**D**) birth year. Legends include respective median (interquartile range) of age and birth years as well as percentage contribution by grouped age categories.

**Supplementary Figure 5.** Percentage distribution of fatal human zoonotic influenza A cases by year of age and birth year, overall and since 2014.

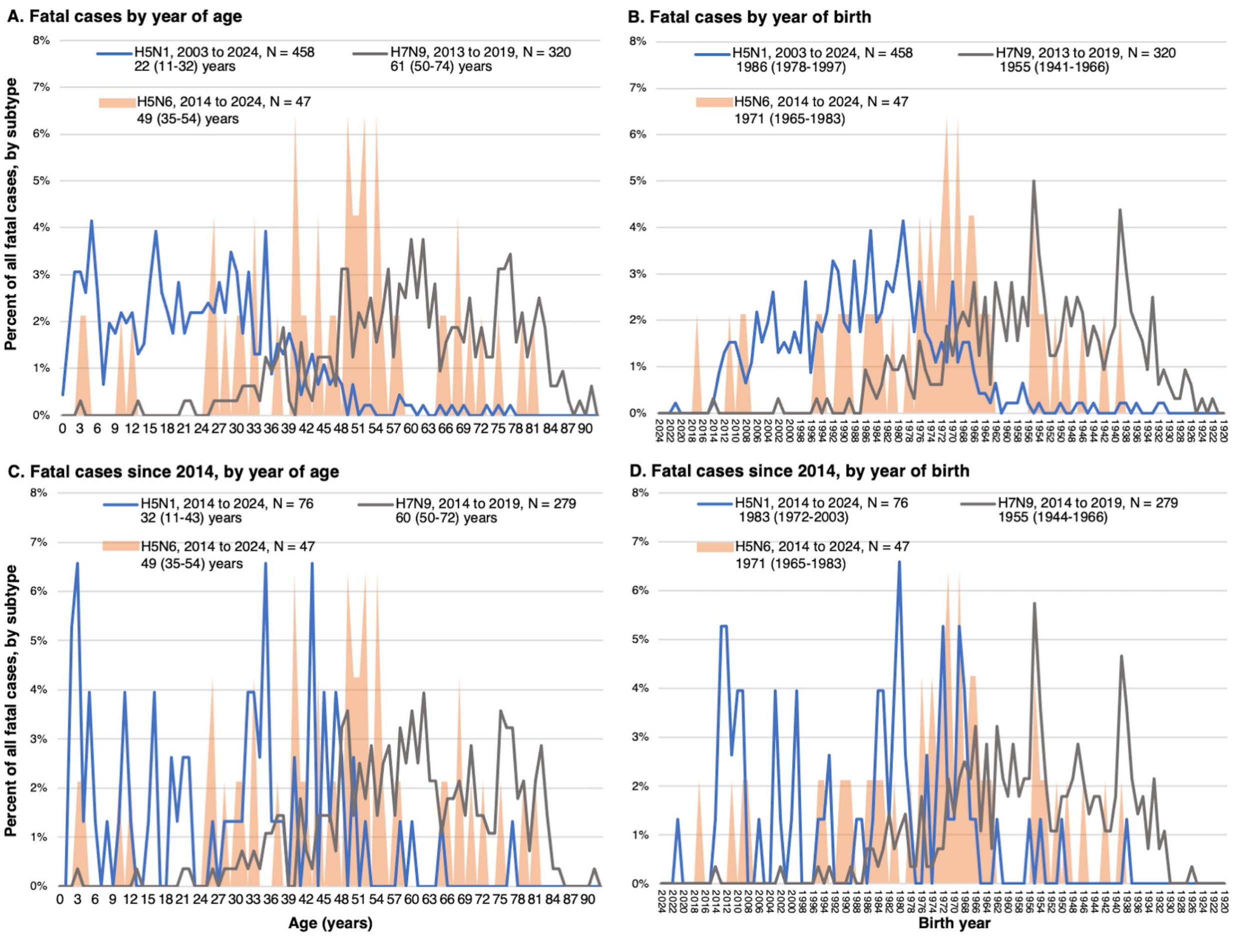


Displayed are the percentage distributions of fatalities due to laboratory-confirmed human zoonotic influenza A H5N1, H5N6 and H7N9 with known age, *overall,* by (**A**) year of age and (**B**) birth year. We display the same information for those with known age *restricted to cases since 2014* by (**C**) year of age; and (**D**) birth year. Legends include respective span of case occurrence years, tallies and median (interquartile range) of age in years or birth years for each zoonotic subtype.

**Supplementary Table 1.** Hemagglutinin (HA) head (HA1): percent amino acid pairwise identities between select influenza A reference viruses.*†

| Influenza A/ strain name | Notes on virus | Avian host H5N1 reference viruses | | | Human zoonotic H5N1 reference viruses | | | | Approved H5N1 vaccine strains for humans | | | | Human adapted H1N1 viruses | | | | | | | | | | | | | | | Non-N1 viruses | | | | | | | |
|---|---|---|---|---|---|---|---|---|---|---|---|---|---|---|---|---|---|---|---|---|---|---|---|---|---|---|---|---|---|---|---|---|---|---|---|
| | | Goose/Guangdong/1/96 | Chicken/Czech_Republic/3744-5/2024 | Duck/Cambodia/W49h3K241D3T/2023 | England/232780677/2023 | Vietnam/KhanhhoaRV1-005/2024 | Texas/37/2024 | British Columbia/PHL-2032/2024 | Vietnam/1194/2004 | Indonesia/5/2005 | Turkey/Turkey/1/2005 | AmericanWigeon/SouthCarolina/22-000345-001/2021 | Washington/001/1918 | Puerto Rico/8/1934 | Fort Monmouth/1/1947 | Kw/1/1957 | USSR/90/1977 | Taiwan/1/1986 | New Caledonia/20/1999 | Solomon Islands/3/2006 | Brisbane/59/2007 | California/07/2009 | Michigan/45/2015 | Brisbane/02/2018 | Guangdong-Maonan/SWI1536/2019 | Victoria/2570/2019 | Victoria/4897/2022 | Rotterdam/1957 | Netherlands/B1/1968 | Hong Kong/001/1968 | Thailand/8/2022 | Changsha/1/2022 | Hong Kong/VM24002346/2024 | Beijing/02/2013 | Gansu/23276/2019 |
| **Avian host H5N1 reference viruses** | | | | | | | | | | | | | | | | | | | | | | | | | | | | | | | | | | | |
| Goose/Guangdong/1/96 | Poultry, clade 0 | – | | | | | | | | | | | | | | | | | | | | | | | | | | | | | | | | | |
| Chicken/Czech_Republic/3744-5/2024 | Poultry, clade 2.3.4.4b | 88 | – | | | | | | | | | | | | | | | | | | | | | | | | | | | | | | | | |
| Duck/Cambodia/W49h3K241D3T/2023 | Poultry, clade 2.3.2.1e | 89 | 88 | – | | | | | | | | | | | | | | | | | | | | | | | | | | | | | | | |
| **Human zoonotic H5N1 reference viruses** | | | | | | | | | | | | | | | | | | | | | | | | | | | | | | | | | | | |
| England/232780677/2023 | Avian-origin Clade 2.3.4.4b | 88 | 99 | 88 | - | | | | | | | | | | | | | | | | | | | | | | | | | | | | | | |
| Vietnam/KhanhhoaRV1-005/2024 | Avian-origin clade 2.3.2.1e | 89 | 87 | 99 | 88 | - | | | | | | | | | | | | | | | | | | | | | | | | | | | | | |
| Texas/37/2024 | Bovine-origin Clade 2.3.4.4b Genotype B3.13 | 87 | 98 | 87 | 98 | 87 | - | | | | | | | | | | | | | | | | | | | | | | | | | | | | |
| British Columbia/PHL-2032/2024 | Avian-origin Clade 2.3.4.4b Genotype D1.1 | 87 | 98 | 87 | 98 | 87 | 97 | - | | | | | | | | | | | | | | | | | | | | | | | | | | | |
| **Approved H5N1 vaccine strains for humans** | | | | | | | | | | | | | | | | | | | | | | | | | | | | | | | | | | | |
| Vietnam/1194/2004 | Clade 1 | 96 | 89 | 90 | 89 | 90 | 88 | 88 | - | | | | | | | | | | | | | | | | | | | | | | | | | | |
| Indonesia/5/2005 | Clade 2.1.3.2 | 95 | 89 | 90 | 89 | 89 | 88 | 88 | 96 | - | | | | | | | | | | | | | | | | | | | | | | | | | |
| Turkey/Turkey/1/2005 | Clade 2.2.1 | 94 | 88 | 91 | 88 | 90 | 87 | 87 | 95 | 94 | - | | | | | | | | | | | | | | | | | | | | | | | | |
| AmericanWigeon/SouthCarolina/22-000345-001/2021 | Clade 2.3.4.4b | 87 | 99 | 87 | 99 | 87 | 99 | 98 | 88 | 88 | 87 | - | | | | | | | | | | | | | | | | | | | | | | | |
| **Human adapted H1N1 viruses** | | | | | | | | | | | | | | | | | | | | | | | | | | | | | | | | | | | |
| Washington/001/1918 | 1918 pandemic (“ASw”) | 58 | 57 | 56 | 57 | 56 | 57 | 56 | 58 | 57 | 57 | 57 | - | | | | | | | | | | | | | | | | | | | | | | |
| Puerto Rico/8/1934 | “A0” | 58 | 58 | 57 | 58 | 57 | 58 | 57 | 58 | 57 | 57 | 58 | 85 | - | | | | | | | | | | | | | | | | | | | | | |
| Fort Monmouth/1/1947 | “Aprime” | 55 | 54 | 54 | 55 | 54 | 54 | 54 | 55 | 54 | 54 | 54 | 84 | 88 | - | | | | | | | | | | | | | | | | | | | | |
| Kw/1/1957 | 1957 H1N1 | 55 | 54 | 54 | 54 | 54 | 54 | 54 | 55 | 53 | 54 | 54 | 82 | 84 | 93 | - | | | | | | | | | | | | | | | | | | | |
| USSR/90/1977 | 1977 pandemic | 53 | 53 | 53 | 54 | 53 | 53 | 53 | 54 | 53 | 53 | 53 | 82 | 87 | 96 | 91 | - | | | | | | | | | | | | | | | | | | |
| Taiwan/1/1986 | Vaccine | 54 | 54 | 54 | 54 | 54 | 54 | 54 | 54 | 53 | 53 | 54 | 82 | 85 | 92 | 90 | 94 | - | | | | | | | | | | | | | | | | | |
| New Caledonia/20/1999 | Vaccine | 55 | 55 | 55 | 55 | 55 | 55 | 55 | 55 | 55 | 54 | 55 | 81 | 84 | 88 | 86 | 88 | 93 | - | | | | | | | | | | | | | | | | |
| Solomon Islands/3/2006 | Vaccine | 55 | 54 | 54 | 54 | 54 | 54 | 54 | 54 | 54 | 53 | 54 | 79 | 82 | 85 | 85 | 85 | 90 | 96 | - | | | | | | | | | | | | | | | |
| Brisbane/59/2007 | Vaccine | 55 | 54 | 55 | 55 | 55 | 55 | 54 | 55 | 55 | 54 | 55 | 79 | 82 | 85 | 84 | 86 | 90 | 97 | 97 | - | | | | | | | | | | | | | | |
| California/07/2009 | 2009 pandemic, vaccine | 56 | 53 | 53 | 54 | 53 | 54 | 53 | 54 | 54 | 54 | 54 | 83 | 76 | 76 | 75 | 74 | 74 | 73 | 73 | 73 | - | | | | | | | | | | | | | |
| Michigan/45/2015 | Vaccine | 56 | 54 | 53 | 55 | 53 | 54 | 54 | 55 | 54 | 54 | 54 | 82 | 75 | 74 | 73 | 72 | 72 | 71 | 72 | 71 | 97 | - | | | | | | | | | | | | |
| Brisbane/02/2018 | Vaccine | 55 | 53 | 53 | 54 | 53 | 53 | 53 | 54 | 53 | 53 | 53 | 81 | 75 | 74 | 73 | 71 | 72 | 72 | 72 | 72 | 94 | 98 | - | | | | | | | | | | | |
| Guangdong-Maonan/SWI1536/2019 | Vaccine | 55 | 53 | 53 | 54 | 53 | 54 | 53 | 54 | 53 | 54 | 54 | 81 | 75 | 74 | 73 | 71 | 72 | 72 | 72 | 72 | 94 | 97 | 97 | - | | | | | | | | | | |
| Victoria/2570/2019 | Vaccine | 56 | 54 | 53 | 55 | 53 | 54 | 54 | 55 | 54 | 54 | 54 | 81 | 75 | 74 | 73 | 71 | 72 | 72 | 72 | 72 | 94 | 97 | 97 | 98 | - | | | | | | | | | |
| Victoria/4897/2022 | Vaccine | 54 | 53 | 53 | 54 | 53 | 53 | 53 | 54 | 53 | 53 | 53 | 81 | 75 | 74 | 73 | 71 | 72 | 72 | 73 | 72 | 91 | 94 | 94 | 96 | 97 | - | | | | | | | | |
| **Non-N1 viruses** | | | | | | | | | | | | | | | | | | | | | | | | | | | | | | | | | | | |
| Rotterdam/1957 | 1957 H2N2 pandemic | 70 | 68 | 68 | 68 | 68 | 67 | 67 | 70 | 70 | 69 | 67 | 61 | 59 | 58 | 58 | 56 | 58 | 58 | 57 | 57 | 55 | 56 | 56 | 56 | 57 | 56 | – | | | | | | | |
| Netherlands/B1/1968 | End of H2N2 epoch | 68 | 66 | 67 | 66 | 67 | 66 | 66 | 68 | 68 | 68 | 66 | 59 | 58 | 56 | 56 | 55 | 56 | 56 | 56 | 55 | 55 | 55 | 55 | 55 | 56 | 55 | 93 | – | | | | | | |
| Hong Kong/001/1968 | 1968 H3N2 pandemic | 37 | 35 | 36 | 35 | 36 | 35 | 35 | 36 | 35 | 35 | 35 | 36 | 34 | 35 | 35 | 34 | 35 | 35 | 36 | 36 | 36 | 37 | 36 | 37 | 38 | 37 | 36 | 36 | – | | | | | |
| Thailand/8/2022 | Recent H3N2 seasonal vaccine | 36 | 34 | 36 | 34 | 36 | 34 | 34 | 36 | 35 | 36 | 34 | 35 | 34 | 33 | 34 | 32 | 33 | 35 | 35 | 35 | 35 | 36 | 36 | 37 | 37 | 36 | 34 | 35 | 80 | – | | | | |
| Changsha/1/2022 | Avian-origin Human zoonotic H5N6 Clade 2.3.4.4b | 88 | 97 | 88 | 97 | 87 | 97 | 97 | 88 | 88 | 87 | 97 | 57 | 58 | 54 | 54 | 53 | 54 | 55 | 55 | 55 | 54 | 55 | 54 | 54 | 55 | 54 | 68 | 66 | 35 | 34 | – | | | |
| Hong Kong/VM24002346/2024 | Avian-origin Human zoonotic H9N2 (LPAI) | 43 | 43 | 43 | 43 | 43 | 43 | 43 | 42 | 42 | 42 | 43 | 46 | 47 | 46 | 47 | 46 | 47 | 47 | 46 | 47 | 44 | 44 | 45 | 45 | 46 | 47 | 47 | 46 | 33 | 33 | 43 | – | | |
| Beijing/02/2013 | Avian-origin Human zoonotic H7N9 (LPAI) | 39 | 38 | 39 | 38 | 39 | 38 | 37 | 39 | 38 | 38 | 38 | 38 | 36 | 38 | 37 | 37 | 37 | 36 | 36 | 36 | 36 | 36 | 35 | 35 | 36 | 36 | 38 | 37 | 36 | 35 | 38 | 35 | – | |
| Gansu/23276/2019 | Avian-origin Human zoonotic H7N9 (HPAI) | 39 | 38 | 38 | 38 | 38 | 38 | 37 | 39 | 39 | 39 | 38 | 38 | 37 | 38 | 38 | 37 | 37 | 36 | 36 | 36 | 36 | 36 | 35 | 35 | 36 | 36 | 36 | 35 | 34 | 34 | 38 | 34 | 93 | – |

LPAI = low pathogenic avian influenza; HPAI = high pathogenic avian influenza. All displayed H5 viruses HPAI.

* Percent amino acid pairwise identities compare 329 amino acids at positions 17-345, H5 numbering according to A/Goose/Guangdong/1/96; one amino acid difference is equal to a 0.3% change in percent identity

† Color gradient according to highest (dark green) and lowest (dark red) percent amino acid identities

**Supplementary Table 2.** Hemagglutinin stalk (HA2): percent amino acid pairwise identities between select influenza A reference viruses.*†

| Influenza A/ strain name | Notes on virus | Avian host H5N1 reference viruses: Goose/Guangdong/196 | Chicken/Czech_Republic/3744-5/2024 | Duck/Cambodia/W49h3K241D3T/2023 | Human zoonotic H5N1 reference viruses: England/232780677/2023 | Vietnam/KhanhhoaRV1-005/2024 | Texas/37/2024 | British Columbia/PHL-2032/2024 | Approved H5N1 vaccine strains for humans: A/Vietnam/1194/2004 | A/Indonesia/5/2005 | A/turkey/Turkey/1/2005 | A/AmericanWigeon/SouthCarolina/22-000345-001/2021 |
|---|---|---|---|---|---|---|---|---|---|---|---|---|
| **Avian host H5N1 reference viruses** | | | | | | | | | | | | |
| Goose/Guangdong/1/96 | Poultry, clade 0 | – | | | | | | | | | | |
| Chicken/Czech_Republic/3744-5/2024 | Poultry, clade 2.3.4.4b | 97 | – | | | | | | | | | |
| Duck/Cambodia/W49h3K241D3T/2023 | Poultry, clade 2.3.2.1e | 95 | 97 | – | | | | | | | | |
| **Human zoonotic H5N1 reference viruses** | | | | | | | | | | | | |
| England/232780677/2023 | Avian-origin Clade 2.3.4.4b | 98 | 100 | 97 | - | | | | | | | |
| Vietnam/KhanhhoaRV1-005/2024 | Avian-origin clade 2.3.2.1e | 96 | 97 | 100 | 98 | - | | | | | | |
| Texas/37/2024 | Bovine-origin Clade 2.3.4.4b Genotype B3.13 | 97 | 98 | 96 | 99 | 97 | - | | | | | |
| British Columbia/PHL-2032/2024 | Avian-origin Clade 2.3.4.4b Genotype D1.1 | 97 | 99 | 97 | 100 | 97 | 98 | - | | | | |
| **Approved H5N1 vaccine strains for humans** | | | | | | | | | | | | |
| A/Vietnam/1194/2004 | Clade 1 | 97 | 99 | 98 | 100 | 98 | 98 | 99 | - | | | |
| A/Indonesia/5/2005 | Clade 2.1.3.2 | 97 | 98 | 96 | 99 | 97 | 98 | 98 | 98 | - | | |
| A/turkey/Turkey/1/2005 | Clade 2.2.1 | 97 | 99 | 97 | 100 | 97 | 98 | 99 | 99 | 98 | - | |
| A/AmericanWigeon/SouthCarolina/22-000345-001/2021 | Clade 2.3.4.4b | 98 | 99 | 97 | 100 | 97 | # | 99 | 99 | 98 | 99 | - |
| **Human adapted H1N1 viruses** | | | | | | | | | | | | |
| Washington/001/1918 | 1918 pandemic ("ASw') | 83 | 83 | 83 | 83 | 83 | 83 | 84 | 83 | 82 | 83 | 83 |
| Puerto Rico/8/1934 | "A0" | 81 | 81 | 81 | 81 | 81 | 80 | 80 | 81 | 80 | 80 | 81 |
| Fort Monmouth/1/1947 | "A prime" | 83 | 82 | 81 | 82 | 82 | 81 | 81 | 82 | 81 | 81 | 82 |
| Kw/1/1957 | 1957 H1N1 | 83 | 82 | 81 | 82 | 81 | 82 | 82 | 82 | 81 | 82 | 82 |
| USSR/90/1977 | 1977 pandemic | 81 | 80 | 81 | 80 | 81 | 80 | 80 | 80 | 79 | 80 | 80 |
| Taiwan/1/1986 | Vaccine | 83 | 82 | 82 | 82 | 83 | 81 | 81 | 82 | 81 | 81 | 82 |
| New Caledonia/20/1999 | Vaccine | 83 | 82 | 83 | 82 | 83 | 82 | 82 | 82 | 81 | 82 | 82 |
| Solomon Islands/3/2006 | Vaccine | 82 | 81 | 82 | 81 | 82 | 81 | 82 | 81 | 80 | 81 | 81 |
| Brisbane/59/2007 | Vaccine | 82 | 81 | 82 | 81 | 82 | 81 | 82 | 81 | 80 | 81 | 81 |
| California/07/2009 | 2009 pandemic, vaccine | 83 | 83 | 83 | 83 | 83 | 82 | 82 | 83 | 82 | 82 | 83 |
| Michigan/45/2015 | Vaccine | 83 | 82 | 82 | 82 | 83 | 82 | 82 | 83 | 81 | 82 | 82 |
| Brisbane/02/2018 | Vaccine | 83 | 82 | 82 | 82 | 83 | 82 | 82 | 83 | 81 | 82 | 82 |
| Guangdong-Maonan/SWl1536/2019 | Vaccine | 83 | 82 | 82 | 82 | 83 | 82 | 82 | 83 | 81 | 82 | 82 |
| Victoria/2570/2019 | Vaccine | 82 | 82 | 82 | 82 | 82 | 81 | 81 | 82 | 81 | 81 | 82 |
| Victoria/4897/2022 | Vaccine | 82 | 82 | 82 | 82 | 82 | 81 | 81 | 82 | 81 | 81 | 82 |
| **Non-N1 viruses** | | | | | | | | | | | | |
| Rotterdam/1957 | 1957 H2N2 pandemic | 89 | 88 | 87 | 88 | 87 | 87 | 88 | 88 | 87 | 87 | 88 |
| Netherlands/B1/1968 | End of H2N2 epoch | 87 | 87 | 86 | 87 | 86 | 87 | 88 | 87 | 86 | 87 | 87 |
| Hong Kong/001/1968 | 1968 H3N2 pandemic | 54 | 54 | 52 | 54 | 52 | 53 | 53 | 54 | 54 | 53 | 54 |
| Thailand/8/2022 | Recent H3N2 seasonal vaccine | 52 | 52 | 50 | 52 | 51 | 51 | 51 | 52 | 52 | 51 | 52 |
| Changsha/1/2022 | Avian-origin Human zoonotic H5N6 Clade 2.3.4.4b | 97 | 99 | 97 | 100 | 97 | 98 | 99 | 99 | 98 | 99 | 99 |
| Hong Kong/VM24002346/2024 | Avian-origin Human zoonotic H9N2 (LPAI) | 62 | 63 | 62 | 63 | 62 | 63 | 64 | 63 | 64 | 63 | 63 |
| Beijing/02/2013 | Avian-origin Human zoonotic H7N9 (LPAI) | 55 | 55 | 54 | 55 | 54 | 55 | 56 | 55 | 55 | 55 | 55 |
| Gansu/23276/2019 | Avian-origin Human zoonotic H7N9 (HPAI) | 55 | 55 | 53 | 55 | 54 | 54 | 55 | 55 | 55 | 54 | 55 |

| Influenza A/ strain name | Human adapted H1N1 viruses: Washington/001/1918 | Puerto Rico/8/1934 | Fort Monmouth/1/1947 | Kw/1/1957 | USSR/90/1977 | Taiwan/1/1986 | New Caledonia/20/1999 | Solomon Islands/3/2006 | Brisbane/59/2007 | California/07/2009 | Michigan/45/2015 | Brisbane/02/2018 | Guangdong-Maonan/SWl1536/2019 | Victoria/2570/2019 | Victoria/4897/2022 |
|---|---|---|---|---|---|---|---|---|---|---|---|---|---|---|---|
| **Human adapted H1N1 viruses** | | | | | | | | | | | | | | | |
| Washington/001/1918 | - | | | | | | | | | | | | | | |
| Puerto Rico/8/1934 | 94 | - | | | | | | | | | | | | | |
| Fort Monmouth/1/1947 | 94 | 96 | - | | | | | | | | | | | | |
| Kw/1/1957 | 92 | 95 | 98 | - | | | | | | | | | | | |
| USSR/90/1977 | 94 | 96 | 98 | 96 | - | | | | | | | | | | |
| Taiwan/1/1986 | 95 | 96 | 98 | 96 | 98 | - | | | | | | | | | |
| New Caledonia/20/1999 | 95 | 96 | 98 | 96 | 98 | 100 | - | | | | | | | | |
| Solomon Islands/3/2006 | 95 | 95 | 97 | 95 | 97 | 98 | 99 | - | | | | | | | |
| Brisbane/59/2007 | 95 | 95 | 97 | 95 | 97 | 98 | 99 | 100 | - | | | | | | |
| California/07/2009 | 92 | 91 | 91 | 90 | 91 | 91 | 92 | 91 | 91 | - | | | | | |
| Michigan/45/2015 | 91 | 91 | 92 | 91 | 91 | 91 | 92 | 91 | 91 | 98 | - | | | | |
| Brisbane/02/2018 | 91 | 91 | 92 | 91 | 91 | 91 | 92 | 91 | 91 | 98 | 100 | - | | | |
| Guangdong-Maonan/SWl1536/2019 | 91 | 91 | 92 | 91 | 91 | 91 | 92 | 91 | 91 | 98 | 100 | 100 | - | | |
| Victoria/2570/2019 | 90 | 90 | 91 | 91 | 90 | 91 | 91 | 90 | 90 | 98 | 100 | 100 | 100 | - | |
| Victoria/4897/2022 | 89 | 89 | 90 | 89 | 89 | 90 | 90 | 89 | 89 | 97 | 98 | 98 | 98 | 98 | - |
| **Non-N1 viruses** | | | | | | | | | | | | | | | |
| Rotterdam/1957 | 84 | 80 | 83 | 83 | 81 | 83 | 83 | 83 | 83 | 81 | 81 | 81 | 81 | 81 | 82 |
| Netherlands/B1/1968 | 82 | 80 | 82 | 83 | 81 | 81 | 82 | 82 | 82 | 81 | 81 | 81 | 81 | 81 | 82 |
| Hong Kong/001/1968 | 57 | 56 | 56 | 56 | 55 | 55 | 55 | 55 | 55 | 58 | 58 | 58 | 58 | 58 | 58 |
| Thailand/8/2022 | 55 | 54 | 54 | 54 | 52 | 53 | 54 | 54 | 54 | 55 | 55 | 55 | 55 | 55 | 55 |
| Changsha/1/2022 | 83 | 80 | 81 | 82 | 80 | 81 | 82 | 81 | 81 | 82 | 82 | 82 | 82 | 81 | 81 |
| Hong Kong/VM24002346/2024 | 64 | 63 | 62 | 63 | 61 | 62 | 63 | 63 | 63 | 62 | 63 | 63 | 63 | 62 | 62 |
| Beijing/02/2013 | 59 | 57 | 57 | 57 | 56 | 56 | 55 | 56 | 56 | 57 | 57 | 57 | 57 | 57 | 58 |
| Gansu/23276/2019 | 58 | 56 | 56 | 56 | 55 | 55 | 54 | 55 | 55 | 56 | 56 | 56 | 56 | 56 | 57 |

| Influenza A/ strain name | Non-N1 viruses: Rotterdam/1957 | Netherlands/B1/1968 | Hong Kong/001/1968 | Thailand/8/2022 | Changsha/1/2022 | Hong Kong/VM24002346/2024 | Beijing/02/2013 | Gansu/23276/2019 |
|---|---|---|---|---|---|---|---|---|
| **Non-N1 viruses** | | | | | | | | |
| Rotterdam/1957 | – | | | | | | | |
| Netherlands/B1/1968 | 98 | – | | | | | | |
| Hong Kong/001/1968 | 54 | 53 | – | | | | | |
| Thailand/8/2022 | 52 | 51 | 93 | – | | | | |
| Changsha/1/2022 | 87 | 87 | 53 | 51 | – | | | |
| Hong Kong/VM24002346/2024 | 62 | 62 | 50 | 49 | 63 | – | | |
| Beijing/02/2013 | 55 | 55 | 66 | 65 | 55 | 57 | – | |
| Gansu/23276/2019 | 54 | 54 | 65 | 65 | 54 | 57 | 97 | – |

LPAI = low pathogenic avian influenza; HPAI = high pathogenic avian influenza. All displayed H5 viruses HPAI.

* Percent amino acid pairwise identities compare 185 amino acids at positions 347-531, H5 numbering according to A/Goose/Guangdong/1/96; one amino acid difference is equal to a 0.5% change in percent identity.

† Color gradient according to highest (dark green) and lowest (dark red) percent amino acid identities

**Supplementary Table 3.** Neuraminidase (NA) head: percent amino acid pairwise identities between select influenza A reference viruses.*†

| Influenza A/ strain name | Notes on virus | Avian host H5N1 reference viruses | Goose/Guangdong/1/96 | Chicken/Czech_Republic/3744-5/2024 | Duck/Cambodia/W49h3K241D3T/2023 | Human zoonotic H5N1 reference viruses | England/232780677/2023 | Vietnam/KhanhhoaRV1-005/2024 | Texas/37/2024 | British Columbia/PHL-2032/2024 | Approved H5N1 vaccine strains for humans | A/Vietnam/1194/2004 | A/Indonesia/5/2005 | A/turkey/Turkey/1/2005 | A/AmericanWigeon/SouthCarolina/22-000345-001/2021 | Select human H1N1 viruses | Washington/001/1918 | Puerto Rico/8/1934 | Fort Monmouth/1/1947 | Kw/1/1957 | USSR/90/1977 | Taiwan/1/1986 | New Caledonia/20/1999 | Solomon Islands/3/2006 | Brisbane/59/2007 | California/07/2009 | Michigan/45/2015 | Brisbane/02/2018 | Guangdong-Maonan/SWL1536/2019 | Victoria/2570/2019 | Victoria/4897/2022 | Non-N1 viruses | Rotterdam/1957 | Netherlands/B1/1968 | Hong Kong/001/1968 | Thailand/8/2022 | Changsha/1/2022 | Hong Kong/VM24002346/2024 | Beijing/02/2013 | Gansu/23276/2019 |
|---|---|---|---|---|---|---|---|---|---|---|---|---|---|---|---|---|---|---|---|---|---|---|---|---|---|---|---|---|---|---|---|---|---|---|---|---|---|---|---|---|
| Avian host H5N1 reference viruses | | | | | | | | | | | | | | | | | | | | | | | | | | | | | | | | | | | | | | | | |
| Goose/Guangdong/1/96 | Poultry, clade 0 | | – | | | | | | | | | | | | | | | | | | | | | | | | | | | | | | | | | | | | | |
| Chicken/Czech_Republic/3744-5/2024 | Poultry, clade 2.3.4.4b | | 95 | – | | | | | | | | | | | | | | | | | | | | | | | | | | | | | | | | | | | | |
| Duck/Cambodia/W49h3K241D3T/2023 | Poultry, clade 2.3.2.1e | | 95 | 94 | – | | | | | | | | | | | | | | | | | | | | | | | | | | | | | | | | | | | |
| Human zoonotic H5N1 reference viruses | | | | | | | | | | | | | | | | | | | | | | | | | | | | | | | | | | | | | | | | |
| England/232780677/2023 | Avian-origin Clade 2.3.4.4b | | 95 | 98 | 93 | | - | | | | | | | | | | | | | | | | | | | | | | | | | | | | | | | | | |
| Vietnam/KhanhhoaRV1-005/2024 | Avian-origin clade 2.3.2.1e | | 95 | 93 | 100 | | 93 | - | | | | | | | | | | | | | | | | | | | | | | | | | | | | | | | | |
| Texas/37/2024 | Bovine-origin Clade 2.3.4.4b Genotype B3.13 | | 95 | 97 | 93 | | 98 | 93 | - | | | | | | | | | | | | | | | | | | | | | | | | | | | | | | | |
| British Columbia/PHL-2032/2024 | Avian-origin Clade 2.3.4.4b Genotype D1.1 | | 94 | 96 | 92 | | 96 | 92 | 96 | - | | | | | | | | | | | | | | | | | | | | | | | | | | | | | | |
| Approved H5N1 vaccine strains for humans | | | | | | | | | | | | | | | | | | | | | | | | | | | | | | | | | | | | | | | | |
| A/Vietnam/1194/2004 | Clade 1 | | 97 | 95 | 95 | | 95 | 95 | 95 | 94 | | - | | | | | | | | | | | | | | | | | | | | | | | | | | | | |
| A/Indonesia/5/2005 | Clade 2.1.3.2 | | 98 | 94 | 95 | | 94 | 95 | 94 | 93 | | 97 | - | | | | | | | | | | | | | | | | | | | | | | | | | | | |
| A/turkey/Turkey/1/2005 | Clade 2.2.1 | | 98 | 95 | 95 | | 95 | 95 | 95 | 94 | | 98 | 97 | - | | | | | | | | | | | | | | | | | | | | | | | | | | |
| A/AmericanWigeon/SouthCarolina/22-000345-001/2021 | Clade 2.3.4.4b | | 96 | 98 | 94 | | 98 | 93 | 99 | 96 | | 95 | 95 | 95 | - | | | | | | | | | | | | | | | | | | | | | | | | | |
| Select human H1N1 viruses | | | | | | | | | | | | | | | | | | | | | | | | | | | | | | | | | | | | | | | | |
| Washington/001/1918 | 1918 pandemic (“ASw’) | | 93 | 94 | 91 | | 94 | 91 | 94 | 94 | | 93 | 91 | 92 | 94 | | - | | | | | | | | | | | | | | | | | | | | | | | |
| Puerto Rico/8/1934 | “A0” | | 88 | 89 | 88 | | 89 | 88 | 89 | 89 | | 88 | 88 | 88 | 89 | | 93 | - | | | | | | | | | | | | | | | | | | | | | | |
| Fort Monmouth/1/1947 | “Aprime” | | 87 | 88 | 87 | | 88 | 87 | 88 | 89 | | 87 | 87 | 87 | 88 | | 92 | 94 | - | | | | | | | | | | | | | | | | | | | | | |
| Kw/1/1957 | 1957 H1N1 | | 86 | 87 | 87 | | 87 | 87 | 87 | 87 | | 87 | 87 | 87 | 87 | | 89 | 92 | 93 | - | | | | | | | | | | | | | | | | | | | | |
| USSR/90/1977 | 1977 pandemic | | 85 | 86 | 86 | | 86 | 86 | 86 | 86 | | 85 | 85 | 86 | 86 | | 87 | 91 | 92 | 95 | - | | | | | | | | | | | | | | | | | | | |
| Taiwan/1/1986 | Vaccine | | 87 | 88 | 87 | | 88 | 87 | 88 | 89 | | 88 | 87 | 88 | 88 | | 90 | 92 | 93 | 95 | 94 | - | | | | | | | | | | | | | | | | | | |
| New Caledonia/20/1999 | Vaccine | | 87 | 88 | 86 | | 88 | 86 | 88 | 88 | | 87 | 87 | 87 | 88 | | 89 | 90 | 90 | 92 | 90 | 93 | - | | | | | | | | | | | | | | | | | |
| Solomon Islands/3/2006 | Vaccine | | 87 | 88 | 86 | | 87 | 86 | 87 | 88 | | 87 | 86 | 87 | 88 | | 89 | 90 | 90 | 92 | 90 | 93 | 99 | - | | | | | | | | | | | | | | | | |
| Brisbane/59/2007 | Vaccine | | 87 | 87 | 86 | | 87 | 85 | 87 | 87 | | 86 | 86 | 87 | 87 | | 89 | 90 | 89 | 91 | 88 | 91 | 98 | 97 | - | | | | | | | | | | | | | | | |
| California/07/2009 | 2009 pandemic, vaccine | | 91 | 92 | 90 | | 91 | 90 | 92 | 91 | | 91 | 90 | 91 | 92 | | 89 | 86 | 86 | 86 | 84 | 86 | 85 | 85 | 85 | - | | | | | | | | | | | | | | |
| Michigan/45/2015 | Vaccine | | 90 | 91 | 89 | | 90 | 89 | 90 | 91 | | 90 | 89 | 90 | 90 | | 89 | 86 | 86 | 85 | 84 | 86 | 85 | 84 | 84 | 97 | - | | | | | | | | | | | | | |
| Brisbane/02/2018 | Vaccine | | 90 | 90 | 89 | | 89 | 89 | 90 | 90 | | 90 | 89 | 90 | 90 | | 88 | 85 | 86 | 85 | 84 | 86 | 84 | 84 | 84 | 97 | 100 | - | | | | | | | | | | | | |
| Guangdong-Maonan/SWL1536/2019 | Vaccine | | 89 | 89 | 89 | | 89 | 88 | 89 | 89 | | 90 | 88 | 89 | 89 | | 88 | 85 | 85 | 85 | 83 | 85 | 84 | 83 | 83 | 96 | 99 | 99 | - | | | | | | | | | | | |
| Victoria/2570/2019 | Vaccine | | 89 | 89 | 88 | | 89 | 88 | 89 | 89 | | 89 | 88 | 89 | 89 | | 88 | 85 | 85 | 85 | 83 | 85 | 84 | 83 | 83 | 96 | 99 | 99 | 100 | - | | | | | | | | | | |
| Victoria/4897/2022 | Vaccine | | 88 | 89 | 88 | | 88 | 88 | 89 | 89 | | 89 | 87 | 88 | 89 | | 87 | 85 | 85 | 84 | 83 | 85 | 84 | 83 | 83 | 95 | 98 | 98 | 99 | 99 | - | | | | | | | | | |
| Non-N1 viruses | | | | | | | | | | | | | | | | | | | | | | | | | | | | | | | | | | | | | | | | |
| Rotterdam/1957 | 1957 H2N2 pandemic | | 45 | 45 | 46 | | 45 | 46 | 45 | 46 | | 46 | 45 | 46 | 45 | | 44 | 44 | 43 | 44 | 44 | 45 | 44 | 43 | 43 | 45 | 45 | 45 | 45 | 45 | 45 | | – | | | | | | | |
| Netherlands/B1/1968 | End of H2N2 epoch | | 45 | 45 | 46 | | 45 | 46 | 45 | 46 | | 46 | 45 | 46 | 45 | | 44 | 44 | 43 | 44 | 44 | 45 | 44 | 44 | 43 | 45 | 45 | 45 | 45 | 45 | 45 | | 95 | – | | | | | | |
| Hong Kong/001/1968 | 1968 H3N2 pandemic | | 45 | 45 | 46 | | 45 | 46 | 45 | 46 | | 46 | 45 | 46 | 45 | | 44 | 44 | 43 | 44 | 44 | 45 | 44 | 44 | 43 | 45 | 45 | 45 | 45 | 45 | 45 | | 95 | 100 | – | | | | | |
| Thailand/8/2022 | Recent H3N2 seasonal vaccine | | 45 | 46 | 46 | | 46 | 46 | 46 | 47 | | 46 | 46 | 46 | 46 | | 45 | 45 | 44 | 45 | 45 | 46 | 45 | 45 | 44 | 45 | 45 | 45 | 45 | 45 | 45 | | 83 | 84 | 84 | – | | | | |
| Changsha/1/2022 | Avian-origin Human zoonotic H5N6 Clade 2.3.4.4b | | 48 | 48 | 48 | | 48 | 49 | 48 | 47 | | 48 | 48 | 48 | 48 | | 47 | 46 | 45 | 46 | 45 | 46 | 44 | 45 | 44 | 47 | 46 | 46 | 46 | 46 | 46 | | 51 | 50 | 50 | 49 | – | | | |
| Hong Kong/VM24002346/2024 | Avian-origin Human zoonotic H9N2 (LPAI) | | 47 | 47 | 48 | | 47 | 48 | 47 | 47 | | 48 | 47 | 48 | 47 | | 45 | 45 | 45 | 46 | 46 | 47 | 46 | 45 | 45 | 46 | 47 | 47 | 47 | 47 | 47 | | 86 | 86 | 86 | 81 | 49 | – | | |
| Beijing/02/2013 | Avian-origin Human zoonotic H7N9 (LPAI) | | 48 | 48 | 48 | | 48 | 48 | 48 | 48 | | 48 | 48 | 48 | 48 | | 47 | 48 | 48 | 47 | 47 | 47 | 46 | 46 | 46 | 48 | 48 | 48 | 47 | 47 | 47 | | 50 | 49 | 49 | 47 | 68 | 49 | – | |
| Gansu/23276/2019 | Avian-origin Human zoonotic H7N9 (HPAI) | | 47 | 47 | 47 | | 47 | 47 | 47 | 47 | | 48 | 47 | 47 | 48 | | 47 | 47 | 47 | 46 | 46 | 47 | 46 | 46 | 45 | 47 | 47 | 47 | 47 | 46 | 46 | | 50 | 49 | 49 | 47 | 67 | 49 | 97 | – |

LPAI = low pathogenic avian influenza; HPAI = high pathogenic avian influenza. All displayed H5 viruses HPAI.

* Percent amino acid pairwise identities compare 388 amino acids at positions 82-469, N1 numbering according to A/California/07/2009; one amino acid difference is equal to a 0.3% change in percent identity

† Color gradient according to highest (dark green) and lowest (dark red) percent amino acid identities

**Supplementary Table 4.** Global distribution of human zoonotic H5N1 cases by country, included in line-list overall* and by period, 1997-2024.

| Country (alphabetical by region) | Overall* N = 964 n (%) | 2003 to 2008 N = 395 (41%) n (%) | 2009 to 2013 N = 255 (26%) n (%) | 2014 to 2023 N = 233 (24%) n (%) | 2024 N = 81 (8%) n (%) |
|---|---|---|---|---|---|
| **Africa** | | | | | |
| Djibouti | 1 | 1 | 0 | 0 | 0 |
| Egypt | 360 (37) | 51 (13) | 123 (48) | 186 (80) | 0 |
| Nigeria | 1 | 1 | 0 | 0 | 0 |
| **Asia** | | | | | |
| Azerbaijan | 8 (1) | 8 (2) | 0 | 0 | 0 |
| Bangladesh | 8 (1) | 1 | 6 (2) | 1 | 0 |
| Cambodia | 72 (7) | 8 (2) | 39 (15) | 15 (6) | 10 (12) |
| China | 56 (6) | 31 (8) | 14 (5) | 10 (4) | 1 (1) |
| India | 1 | 0 | 0 | 1 | 0 |
| Indonesia | 200 (21) | 141 (36) | 54 (21) | 5 (2) | 0 |
| Iraq | 3 | 3 (1) | 0 | 0 | 0 |
| Laos | 3 | 2 (1) | 0 | 1 | 0 |
| Myanmar | 1 | 1 | 0 | 0 | 0 |
| Nepal | 1 | 0 | 0 | 1 | 0 |
| Pakistan | 3 | 3 (1) | 0 | 0 | 0 |
| Thailand | 25 (3) | 25 (6) | 0 | 0 | 0 |
| Turkey | 12 (1) | 12 (3) | 0 | 0 | 0 |
| Vietnam | 129 (13) | 107 (27) | 18 (7) | 3 (1) | 2 (2) |
| **Australia** | | | | | |
| Australia | 1 | 0 | 0 | 0 | 1 (1) |
| **Europe** | | | | | |
| Spain | 2 | 0 | 0 | 2 (1) | 0 |
| United Kingdom | 5 (1) | 0 | 0 | 5 (2) | 0 |
| **North America** | | | | | |
| Canada | 2 | 0 | 1 | 0 | 1 (1) |
| United States | 67 (7) | 0 | 0 | 1 | 66 (81) |
| **South America** | | | | | |
| Chile | 1 | 0 | 0 | 1 | 0 |
| Ecuador | 1 | 0 | 0 | 1 | 0 |

Tallies of human zoonotic influenza A cases due to H5N1 by country and period, included in our assembled H5N1 line-list spanning occurrence dates between 2003 and December 31, 2024. Countries are organized alphabetically by region. Tallies without accompanying percentages represent <1% of all cases by country and period. See Supplementary Note 1 for details related to dataset completeness.

* As per World Health Organization cumulative tallies (through December 12, 2024) (*98*), the 18 cases from China in 1997 are not included in our dataset, otherwise available in line-list from Lai et al, 2016[10], and Chan, 2002[11].

**Supplementary Table 5.** Neuraminidase (NA) and hemagglutinin (HA) sequences of reference viruses used in pairwise identity comparisons obtained from the Global Initiative on Sharing All Influenza Data (GISAID).

| NA Segment ID | HA Segment ID | Country | Collection date | Isolate-ID | Isolate name | Originating Lab | Submitting Lab | Authors |
|---|---|---|---|---|---|---|---|---|
| EPI5787 | EPI5797 | China | 1996 | EPI_ISL_1254 | A/Goose/Guangdong/1/96 | Centres for Disease Control and Prevention | Centres for Disease Control and Prevention | Xu,X, Subbarao, Cox,N.J., Guo,Y. |
| EPI3139831 | EPI3139832 | Czech Republic | 2024-Feb-26 | EPI_ISL_19000410 | A/chicken/Czech_republic/3744-5/2024 | State Veterinary Institute Prague | State Veterinary Institute Prague | Nagy,A;Cernikova,L;Klickova,E; Sterbova,M;Orenicova,K |
| EPI3169189 | EPI3169190 | Cambodia | 2023-Dec-08 | EPI_ISL_19025576 | A/Duck/Cambodia/W49h3K241D3T/2023 | Institut Pasteur du Cambodia | Institut Pasteur du Cambodia | Yann,S;Kol,S;Siegers,J;Horm,SV;Tum,S;San,S;Duong,V;Karlsson,E |
| EPI2718822 | EPI2718820 | United Kingdom | 2023-Jul-07 | EPI_ISL_18161874 | A/England/232780677/2023 | UK Health Security Agency – Colindale | UKHSA/Respiratory Virus Unit | UKHSA, Respiratory Virus Unit |
| EPI3181130 | EPI3181128 | Vietnam | 2024-Mar-19 | EPI_ISL_19031556 | A/Vietnam/KhanhhoaRV1-005/2024 | Pasteur Institute in Nha Trang | National Institute of Hygiene and Epidemiology (NIHE) | Hung,D T; Anh,D T;Anh,H T |
| EPI3171486 | EPI3171488 | United States | 2024-Mar-28 | EPI_ISL_19027114 | A/Texas/37/2024 | Texas Department of State Health Services-Laboratory Services | Centers for Disease Control and Prevention | Presley, S M; Webb, Cynthia R; Malaeb, S; Cofi, M; Davis, T; Kondor, B; Steel, J; Kirby, M; Sheffield, S; Liddell, J; Frederick, J; Di, H; Pusch, E; Lacek, K; Barnes, J |
| EPI3650016 | EPI3650014 | Canada | 2024-Nov-09 | EPI_ISL_19548836 | A/British_Columbia/PHL-2032/2024 | British Columbia Centre for Disease Control (BCCDC); Public Health Agency of Canada (PHAC) | B.C. Centre for Disease Control | Shannon Russell, Natalie Prystajecky, Linda Hoang, Agatha Jassem, James Zlosnik, John Tyson, Frankie Tsang, Jessica Caleta, John Palmer, Dan Fornika, Kevin Yang, Kevin Kuchinski, Tracy Lee, Rob Azana, Janet Fung, Michael Chan, Branco Cheung, Nathalie Bastien, Ruimin Gao, Cody Buchanan, Jasmine Frost, Taeyo Chestley, Charlene Ranadheera |
| EPI1256206 | EPI1256205 | Vietnam | 2018-Jul-10 | EPI_ISL_314984 | A/Vietnam/1194/2004_NIBRG-14_(18/136) | | National Institute for Biological Standards and Control (NIBSC) | Nicolson, C |
| EPI376539 | EPI376537 | Indonesia | 2005 | EPI_ISL_121541 | A/Indonesia/5/2005 | Erasmus Medical Center | Erasmus Medical Center | Herfst,S., Schrauwen,E.J., Linster,M, Chutinimitkul,S., de Wit,E., Munster,V.J., Sorrell,E.M, Bestebroer,T.M, Burke,D.F., Smith,D.J., Rimmelzwaan,G.F., Osterhaus,A.D. and Fouchier,R.A. |
| EPI118777 | EPI118794 | Turkey | 2005 | EPI_ISL_10107 | A/turkey/Turkey/1/2005 | | | |
| EPI2709127 | EPI2709137 | United States | 2021-Dec-30 | EPI_ISL_18133029 | A/American Wigeon/South Carolina/22-000345-001/2021 | | | Youk,S., Torchetti,M.K., Lantz,K., Lenoch,J.B., Killian,M.L., Leyson,C., Bevins,S.N., Dilione,K., Ip,H.S., Stallknecht,D.E., Poulson,R.L., Suarez,D.L., Swayne,D.E.; Pantin-Jackwood,M.J. |
| EPI2971421 | EPI2971423 | United States | 1918-Sept-29 | EPI_ISL_18846999 | A/Washington/NIAID-001/1918 | National Institute of Allergy and Infectious Diseases | National Institute of Allergy and Infectious Diseases | Xiao, Y |
| EPI252286 | EPI252235 | Puerto Rico | 1934 | EPI_ISL_73427 | A/Puerto_Rico/8/34/Mount_Sinai | | | Schickli,J.H.; Flandorfer,A.; Nakaya,T.; Martinez-Sobrido,L.; Garcia-Sastre,A.; Palese,P. |
| EPI30341 | EPI30336 | United States | 1947 | EPI_ISL_5169 | A/FortMonmouth/1/1947 | | | |
| EPI400031 | EPI400029 | China | 1957 | EPI_ISL_130407 | A/Kw/1/1957 | | | Wentworth,D.E.; Dugan,V.; Halpin,R.; Lin,X; Bera,J.; Wester,E.; Ghedin,E.; Fedorova,N.; Tsitrin,T.; McLellan,M; Stockwell,T.; Amedeo,P.; Appalla,L.; Bishop,B.; Edworthy,P.; Gupta,N.; Hoover,J.; Katzel,D.; Li,K.; Schobel,S.; Shrivastava,S.; Thovarai,V.; Wang,S.; Webster,R.; |

| NA Segment ID | HA Segment ID | Country | Collection date | Isolate-ID | Isolate name | Originating Lab | Submitting Lab | Authors |
|---|---|---|---|---|---|---|---|---|
| | | | | | | | | Webby,R.; Krauss,S.; Bao,Y.; Sanders,R.; Dernovoy,D.; Kiryutin,B.; Lipman,D.J.; Tatusova,T. |
| EPI241094 | EPI241092 | Russian Federation | 1977 | EPI_ISL_69311 | A/USSR/90/1977 | | | Cummings,I.W.; Salser,W.A. |
| EPI105058 | EPI105054 | Taiwan | 1986 | EPI_ISL_10251 | A/Taiwan/1/1986 | | | |
| EPI163941 | EPI105019 | New Caledonia | 1999 | EPI_ISL_649 | A/New Caledonia/20/1999 | | | |
| EPI509400 | EPI509399 | Solomon Islands | 2006-Aug-21 | EPI_ISL_157458 | A/Solomon Islands/3/2006 | WHO Centre for Reference &Research on Influenza | National Institute for Medical Research | |
| EPI457184 | EPI457183 | Australia | 2007-Jul-01 | EPI_ISL_142730 | A/Brisbane/59/2007 | | | Sabaiduc,S.; Petric,M; Skowronski,D.M |
| EPI185379 | EPI177294 | United States | 2009-Apr-09 | EPI_ISL_31553 | A/California/07/2009 | Naval Health Research Center (NHRC) U.S. Navy | Centers for Disease Control and Prevention | |
| EPI662593 | EPI662594 | United States | 2015-Sept-07 | EPI_ISL_199532 | A/Michigan/45/2015 | Michigan Department of Community Health | Centers for Disease Control and Prevention | |
| EPI212885 | EPI1212884 | Australia | 2018-Jan-04 | EPI_ISL_306350 | A/Brisbane/02/2018 | Queensland Health Forensic and Scientific Services | WHO Collaborating Centre for Reference and Research on Influenza | Dneg,Y-M; Iannello,P.; Lau,H.; Kaye,M; Todd,A.; Spirason,N.; Komadina,N. |
| EPI 1542569 | EPI1542570 | China | 2019-Jun-17 | EPI_ISL_377080 | A/Guangdong-Maonan/SWL1536/2019 | WHO Chinese National Influenza Center | WHO Chinese National Influenza Center | Xiaoxu,Zeng;Xiyan,Li;Weijuan,Huang; Lei,Yang;Dayan,Wang |
| EPI2222757 | EPI2222758 | Australia | 2019-Nov-22 | EPI_ISL_15907684 | A/Victoria/2570/2019 | Alfred Hospital | WHO Collaborating Centre for Reference and Research on Influenza | Deng,Y-M; Barr,I; Aziz,A |
| EPI2235968 | EPI2235969 | | 2022-Oct-02 | EPI_ISL_16003490 | A/Victoria/4897/2022 | Royal Melbourne Hospital | | |
| EPI297877 | EPI297875 | Netherlands | 1957 | EPI_ISL_84911 | A/Rotterdam/1957 | | | Fouchier, R |
| EPI542508 | EPI542494 | Netherlands | 1968-Feb-29 | EPI_ISL_166563 | A/Netherlands/B1/1968 | | | Linster,M; Lexmond,P.; Bestebroer,T.; Osterhaus,A.; Fouchier,R.; Herfst,S. |
| EPI2595385 | EPI2595399 | Hong Kong (SAR) | 1968-Jan-01 | EPI_ISL_17805317 | A/Hong Kong/001/1968 | | | Chen,P.; Jin,Z.; Peng,L.; Zheng,Z.; Cheung,Y.M; Guan,J.; Chen,L.; Huang,Y.; Fan,X; Zhang,Z.; Shi,D.; Xie,J.; Chen,R.; Xiao,B.; Yip,C.H.; Holmes,E.C.; Lam,T.Y.; Zhu,H.; Guan,Y. |
| EPI2384159 | EPI2384160 | Thailand | 2022-Jul-11 | EPI_ISL_16864398 | A/Thailand/8/2022 | WHO National Influenza Centre, National Institute of Medical Research (NIMR) | WHO Collaborating Centre for Reference and Research on Influenza | Okada,P; Yuigun,S; Kala,S; Deng,Y-M; Barr,I; Aziz,A |
| EPI2287050 | EPI2287052 | China | 2022-Nov-11 | EPI_ISL_16466440 | A/Changsha/1/2022 | Changsha center for diease control and prevetion | Changsha Disease Prevention and Control Center | Huang, Z; Xinhua, O |
| EPI509073 | EPI509071 | China | 2013-May-27 | EPI_ISL_157286 | A/Beijing/02/2013 | | WHO Chinese National Influenza Centre | Wang,Dayan; Gao,Rongbao; Yang,Lei; Li,Xiyan; Cheng,Yanhui; Zou,Shumei; Zhou,Jianfang; Zhu,Wenfei; Guo,Junfeng; Dong,Jie; Zhang,Ye; Liu,Liqi; Bo,Hong; Xin,Li; Huang,Weijuan; Dong,Libo; Zhao,Xiang; Lan,Yu; Li,Xiaodan; Shu,Yuelon |
| EPI1431599 | EPI1431600 | China | 2019-Apr-03 | EPI_ISL_353998 | A/Gansu/232762019 | Gansu Provincial Centre for Disease Control and Prevention | | Wang, Dayan; Yu, Deshan; Liu, Jia; Li, Xiyan |
| EPI3059194 | EPI3059195 | Hong Kong (SAR) | 2024-Feb-16 | EPI_ISL_18926219 | A/Hong Kong/VM24002346/2024 | Hong Kong Department of Health | Hong Kong Department of Health | Mak, Gannon.C.K; Tsang, Alan.K.L; Lo, Janice.Y.C. |

**Supplementary Information, References**


1. World Health Organization. Cumulative number of confirmed human cases for avian influenza A(H5N1) reported to WHO, 2003-2024,12. https://www.who.int/publications/m/item/cumulative-number-of-confirmed-human-cases-for-avian-influenza-a(h5n1)-reported-to-who--2003-2024--20-december-2024 (2024).
2. Global Influenza Programme, World Health Organization. Risk assessments and summaries of influenza at the human-animal interface, monthly. https://www.who.int/teams/global-influenza-programme/avian-influenza/monthly-risk-assessment-summary (2024).
3. World Health Organization. Disease Outbreak News. https://www.who.int/emergencies/disease-outbreak-news (2024).
4. World Health Organization. H5N1 highly pathogenic avian influenza: Timeline of major events. https://cdn.who.int/media/docs/default-source/influenza/avian-and-other-zoonotic-influenza/h5n1_avian_influenza_update20141204.pdf?sfvrsn=d1846969_5&download=true (2014).
5. Western Pacific Regional Office, World Health Organization. Avian influenza. https://www.who.int/westernpacific/wpro-emergencies/surveillance/avian-influenza (2024).
6. University of Minnesota. *Center for Infectious Disease Research and Policy (CIDRAP).*
7. International Society for Infectious Diseases. Program for Monitoring Emerging Diseases (ProMED). https://www.promedmail.org/.
8. FluTrackers. https://flutrackers.com/.
9. Fiebig, L. *et al.* Avian influenza A(H5N1) in humans: new insights from a line list of World Health Organization confirmed cases, September 2006 to August 2010. *Euro Surveill* **16**, (2011).
